# Advancing Human Population Genomics with DNA Foundation Models

**DOI:** 10.1101/2025.09.09.25335438

**Authors:** Rui Zhu, Xiaopu Zhou, Marla Mendes de Aquino, Worrawat Engchuan, Hang Zhou, Haoyu Cheng, Alzheimer’s Disease Neuroimaging Initiative, John Hardy, Haixu Tang, Stephen W. Scherer, Lucila Ohno-Machado

**Author notes:** Rui Zhu and Xiaopu Zhou contributed equally to this work. Part of the data used in preparation of this article were obtained from the Alzheimer’s Disease Neuroimaging Initiative (ADNI) database (adni.loni.usc.edu). As such, the investigators within the ADNI contributed to the design and implementation of the ADNI and/or provided data but did not participate in analysis or writing of this report. A complete listing of ADNI investigators can be found at http://adni.loni.usc.edu/wp-content/uploads/how_to_apply/ADNI_Acknowledgement_List.pdf. Corresponding authors: Correspondence to: Xiaopu Zhou and Lucila Ohno-Machado.

## Abstract

DNA foundation models offer a new approach to interpret genetic variation, but their potential in population-scale genomics remains untapped. We introduce a novel analytical framework that integrates a genomic foundation model with human population genomics studies. We employed the Evo2 DNA foundation model to systematically score the functional impact of a variant and haplotype across diverse cohorts including people with Alzheimer’s Disease Neuroimaging Initiative (ADNI), the Human Pangenome Project, and the UK Biobank. As proof-of-concept, the analysis of the *APOE* locus confirmed the approach’s validity, with model-derived scores can help to prioritize putatively functional variants and quantify effects of both variants and haplotype onto Alzheimer’s Disease susceptibility or associated endophenotypes, including cognitive performance, brain structural change and amyloid load. Specifically, scoring multi-ancestry assembly sequences from the Human Pangenome Project revealed, for the first time, that genetic variation could nicely explains ancestry-specific differences in APOE expression and the impact of *APOE*-ε4 on Alzheimer’s disease risk. Overall, this study provides a scalable framework for mapping functional genetic variation, complementing conventional population-genomics approaches, enabling better interpretation of genetic effects in complex genomic regions at population scale.

## Main Text

Foundation models (*1–4*), large-scale neural networks trained on diverse data, have revolutionized natural language and vision tasks and now promise to have similar impact in genomics. Newly emerging DNA foundation models, including Evo and Evo2 (*5*, *6*), learn the “language” of the genome from sequences spanning the tree of life allowing the prediction of putative functional effects from sequence context. While these models have been used primarily for reference genome annotation (*7*, *8*), their potential for population-scale discovery is largely untapped.

Meanwhile, biobank-scale resources (*9*, *10*) linking whole-genome sequences to rich phenotypes now encompass millions of individuals. Even studies of this magnitude often lack the statistical power to pinpoint causal variants underlying rare but high-mortality diseases, and in common disorders like Alzheimer’s disease (AD), extensive linkage disequilibrium yields complex haplotypes that defy exhaustive association testing (*11–13*). Although whole-genome sequencing provides a comprehensive catalog of variants, extracting mechanistic insight from these data demands new analytical approaches capable of inferring functional impact directly from sequence.

These challenges are exemplified by the *APOE* locus, the strongest genetic risk factor for AD, a neurodegenerative disease affecting ∼10% of people over age 65. From an evolutionary standpoint, the three coding *APOE* alleles ε2, ε3, and ε4, represent a fascinating example of balancing selection, genetic drift, and life-history trade-offs. Each has distinct origins, functions, and consequences for human health and aging(*14*, *15*). Two alleles, ε4 and ε2, strongly modulate AD susceptibility: possession of ancestral allele ε4 confers roughly a 3-15-fold higher risk, whereas derived allele ε2 (from ∼80,000 years ago) is partially protective (*16*, *17*). The common allele ε3 (found in 80% in general population and was derived between 200,000 and 300,000 years ago) has neutral effect on AD risk (*15*). Critically, ε4’s penetrance is much higher in European ancestries than in African, underscoring the influence of local ancestry genomic context (*18–20*). Prior fine-mapping (*13*) of this region identified additional noncoding risk signals and delineated an ∼80-kb ε4-harboring haplotype spanning *APOE* and neighboring genes (*NECTIN2*, *TOMM40*, *APOC1*). However, traditional methods resolved only the most common haplotypes, precluding precise individualized risk estimates (*12*, *13*, *21*, *22*).

Now, with the advent of large biobanks and disease-specific cohorts (e.g., ADNI (*23*)), alongside Human Pangenome Project (*24*) delivering an unprecedented, ancestry-spanning atlas of genetic variation, there is opportunity to deploy sequence-based DNA foundation models on population genomic data. An overview of our study design is shown in **Fig. 1**. Evo 2, in particular, can analyze up to 1 megabase (Mb) of DNA sequence efficiently, enabling evaluation of a wide range of variants—including single nucleotide polymorphisms (SNPs), insertions/deletions, structural variations, and complex haplotypes in a zero-shot manner (without task-specific fine-tuning). Building on Evo 2-7B (*25*), we present a new analytical framework that integrates a DNA foundation model with biobank whole-genome sequences, ancestry-specific reference panels, and genome-wide association study (GWAS) summary statistics to generate quantitative haplotype-level risk maps. As a proof of concept, we applied this framework to the well-characterized *APOE* locus (*18*, *13*) implicated in AD. This approach pinpointed functionally important genetic variants and haplotypes across individuals and populations, highlighting the powerful synergy between modern AI foundation models and population-scale genomics.

**Figure 1.**
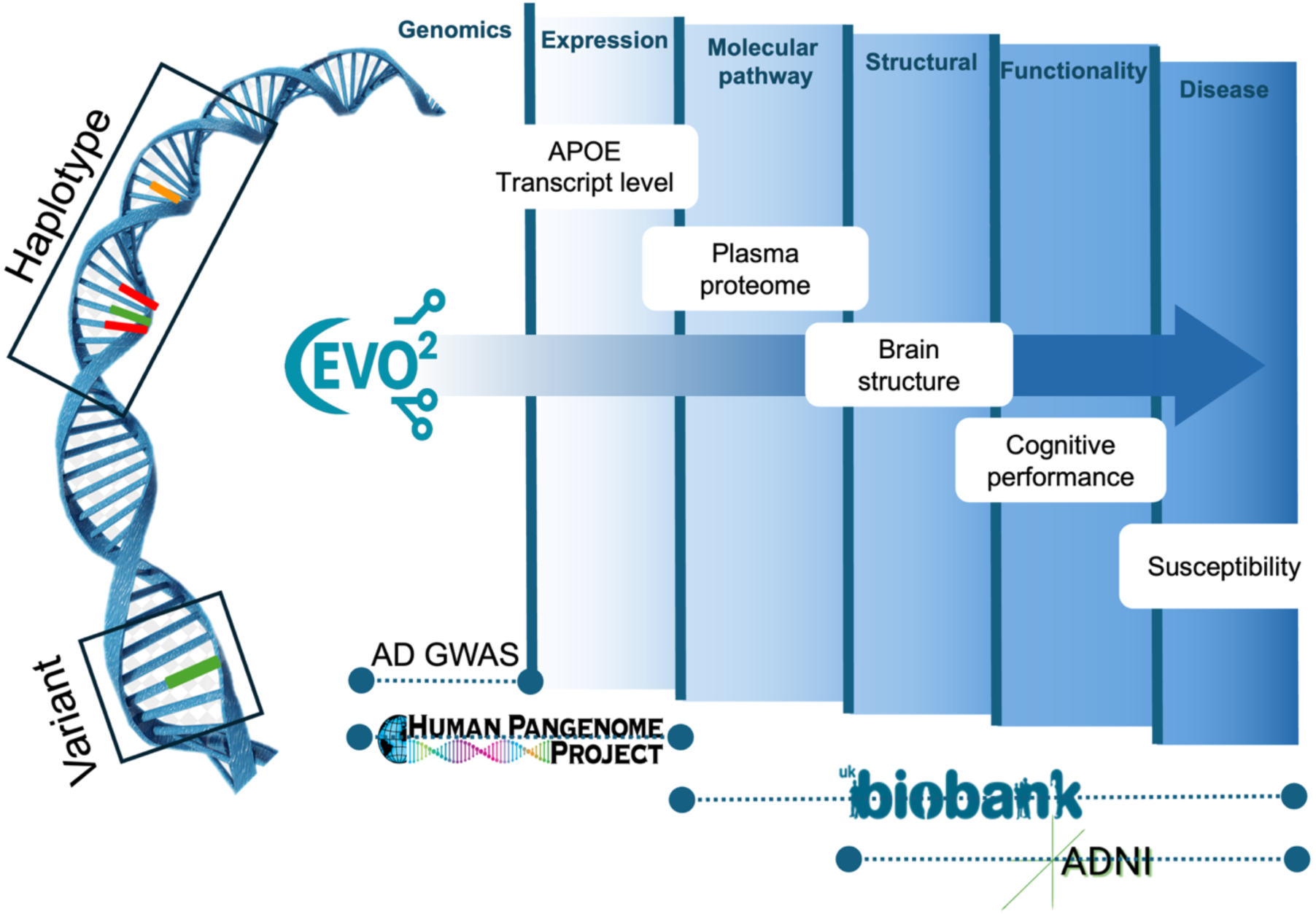
Study Design. A conceptual workflow for applying a DNA foundation model (Evo2) to population genomics. The approach performs variant- and haplotype-level analyses and evaluates how DNA foundation model-derived functional scores associate with multi-layered biological data—from genomic variation and gene expression to molecular pathways, brain structure, cognitive function, and disease susceptibility. The framework integrates data from large-scale population resources, including GWAS for Alzheimer’s disease (AD), the Human Pangenome Project, UK Biobank, and the Alzheimer’s Disease Neuroimaging Initiative (ADNI), to connect sequence-derived functional insights to complex phenotypes in population cohorts. The APOE locus serves as a proof-of-concept example, demonstrating how sequence-based functional predictions can be linked to AD-related phenotypes.

To evaluate the applicability of the Evo2 in prioritizing genetic risk at the single-variant level for disease susceptibility, we analyzed GWAS data for AD reported by Jansen et al (*26*). (**Table S1**). We included variants annotated with dbSNP v156 rsIDs, a minor allele frequency (MAF) greater than 5%, and that passed both a nominal p-value threshold of 1×10⁻⁴ and a false discovery rate (FDR) threshold of 0.05 within the 80-kb window spanning *APOE* region and its surrounding locus (chr19:45,345,000–45,425,000; GRCh37). A total of 211 variants met these criteria and were retained for subsequent validation (**Table S2**).

### Evo2 scores explain variants’ contributions to Alzheimer’s disease risk

We obtained the Evo2 variant score (denoted as the Evo2 score) by calculating the contrast value between 80-kb DNA fragment windows with and without the non-reference allele (see **Supplementary A** and **Fig. S1–S2** for methodological details). Interestingly, although multiple variants in the studied locus showed strong associations with Alzheimer’s disease (as indicated by large absolute z-scores from the AD GWAS; **Fig. 2A**), Evo2 successfully prioritized the *APOE*-ε2 (rs7412) and *APOE*-ε4 (rs429158) alleles as the top variants exerting the greatest biological impact among those selected. Interestingly, the derived and protective allele *APOE*-ε2 exhibits negative Evo2 score (−24.20; likely representing a more evolutionarily novel event; **Table S2**), while the ancestral and risk allele *APOE*-ε4 has positive Evo2 score (15.29; likely representing a more evolutionarily familiar event; **Table S2**). This pattern shows that, at this locus, Evo2 scores somehow correlate with individual variant risk effect toward AD, with a higher Evo2 score can coincide with greater susceptibility to AD. In addition, several variants in the surrounding *APOE* region were also highlighted by the Evo2 score, corroborating our previous findings that noncoding variants contribute to *APOE*-associated risk (**Fig. 2A**)(*13*). Further analysis revealed a significant correlation between the Evo2 score and both AD association significance (Z-score) and effect size (beta coefficient) among all 211 AD-associated variants (Pearson r ≈ 0.14, p < 0.05; **Fig. 2B**). This correlation was stronger among a subset of variants that showed low linkage disequilibrium with one another (Pearson r ≈ 0.40, p < 0.05; **Fig. 2C**).

**Figure 2.**
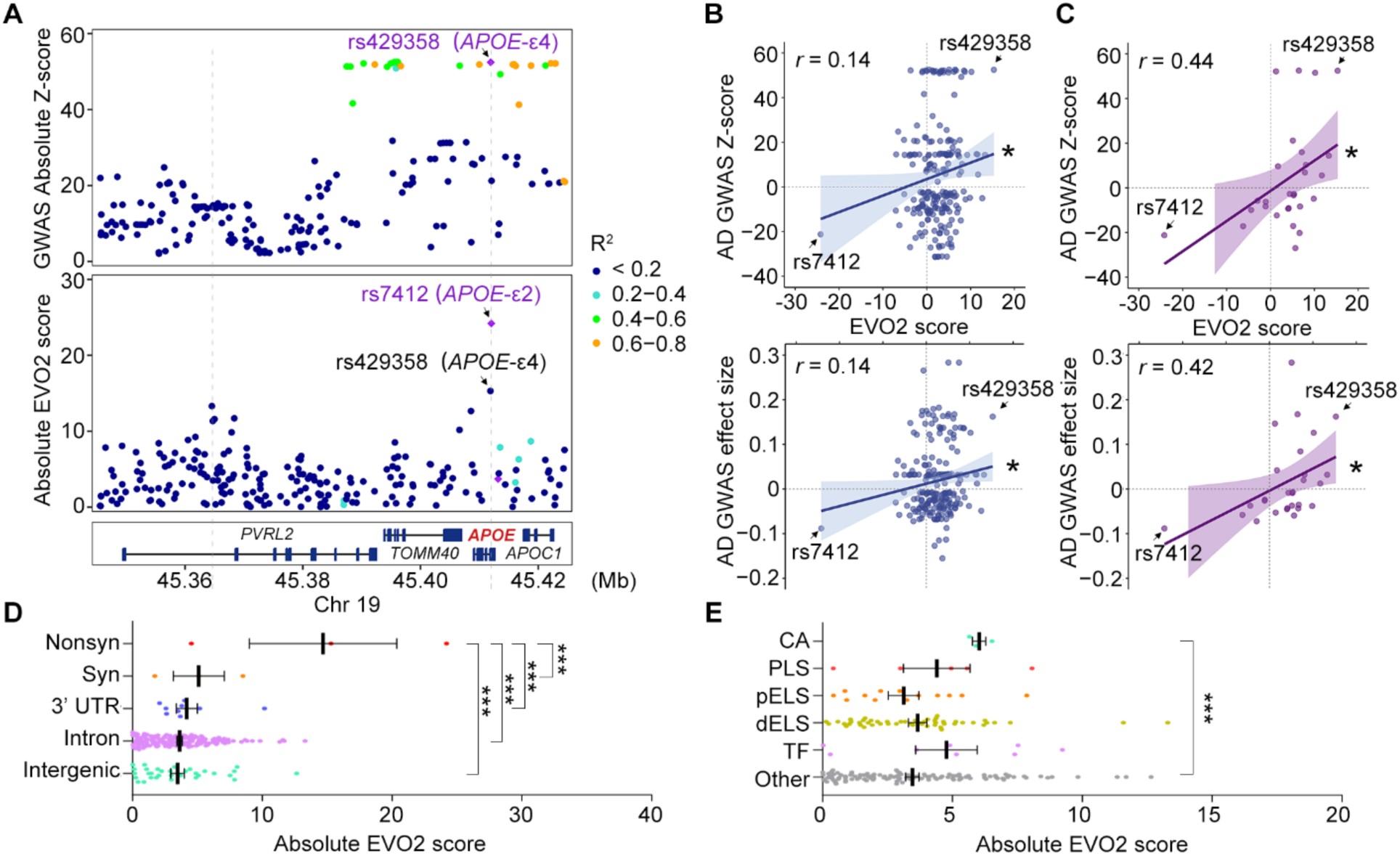
EVO2 Scores at the APOE locus Mirror GWAS Signals and Effect Sizes in AD GWAS Data. **(A)** Regional plot showing selected Alzheimer’s disease (AD)-associated variants within the *APOE* locus and surrounding regions. The upper panel displays absolute GWAS z-scores, and the lower panel shows the corresponding absolute Evo2 scores. Key coding variants (*rs429358* and *rs7412*) are highlighted. **(B–C)** Correlation analyses between Evo2 scores and **(B)** absolute GWAS z-scores and variant effect sizes for all selected variants (n = 211), and **(C)** a subset of variants selected via LD clumping (n = 28). Pearson correlation test; *p* < 0.05. **(D–E)** Comparison of absolute Evo2 scores among stratified variant groups for **(D)** all variants and **(E)** noncoding variants. One-way ANOVA with Benjamini–Krieger–Yekutieli correction (two-stage linear step-up procedure); \*\**p* < 0.001. 3′ UTR, 3′ untranslated region; AD, Alzheimer’s disease; CA, chromatin accessibility; dELS, distal enhancer-like signature; GWAS, genome-wide association study; Intergenic, Intergenic region; Intron, intronic region; LD, linkage disequilibrium; Nonsyn, nonsynonymous variant; Other, other regulatory annotations; pELS, proximal enhancer-like signature; PLS, promoter-like signature; Syn, synonymous variant; TF, transcription factor binding site.

We further examined the distribution of Evo2 scores across variant groups stratified by their biological properties. The results showed that nonsynonymous variants (those altering amino acids) had significantly higher Evo2 scores compared to other variant types (p < 0.05; **Fig. 2D**). Additionally, there was a trend toward higher Evo2 scores for variants located in exonic regions, such as synonymous variants and those in the 3′ UTR, compared to variants located in intronic or intergenic regions (**Fig. 2D**). We conducted a similar analysis focusing on variants in noncoding regions. The results revealed that variants residing in regions of active chromatin had significantly higher Evo2 scores compared to those outside annotated regulatory regions (p < 0.05; **Fig. 2E**).

Overall, these findings established the sequence-based Evo2 score as quantitative measure of a variant’s impact. By effectively capturing functional signals, the score successfully explains known variant associations within the *APOE* locus and its surrounding regions.

### Evo2 scores capture haplotype effects across diverse population backgrounds

Previous research (*13*) has identified local haplotypes in this region that modulate AD risk. We therefore hypothesized that the Evo2 framework could capture these aggregate, haplotype-level effects. By calculating Evo2 scores for haplotypes carrying multiple AD-associated variants, we aim to model the joint impact of those variants. Extending our single-variant analysis, we generated an Evo2 haplotype score by replacing each phased 80-kb haplotype in the model and comparing its log-likelihood with that of the corresponding GRCh38 reference sequence (see **Supplementary Information** for detailed methods).

We first applied this haplotype-based score to the 240 high-quality, fully phased genomes from the Human Pangenome Reference Consortium to test whether Evo2 captures local haplotype influences on disease susceptibility. Interestingly, Evo2 haplotype scores track APOE expression across populations: for ancestries with higher mean Evo2 haplotypes scores, they displayed higher mean APOE mRNA levels in lymphoblastoid cell lines (Pearson r = 0.86, P < 2 × 10⁻¹⁶; **Fig. 3A**; **Table S3**). The pattern persisted at the individual level. In the 33 participants with both genome assemblies (haplotype sequences) and expression data, the score of the lower-scoring (“alternative”) haplotype rose in step with transcript abundance (P < 0.05), whereas the higher-scoring partner exerted no detectable effect (**Fig. 3B**; **Table S4**). Thus, Evo2 haplotype scoring can stratify haplotypes by their regulatory impact and recapitulates transcriptional variation, despite being trained without any expression data.

**Figure 3.**
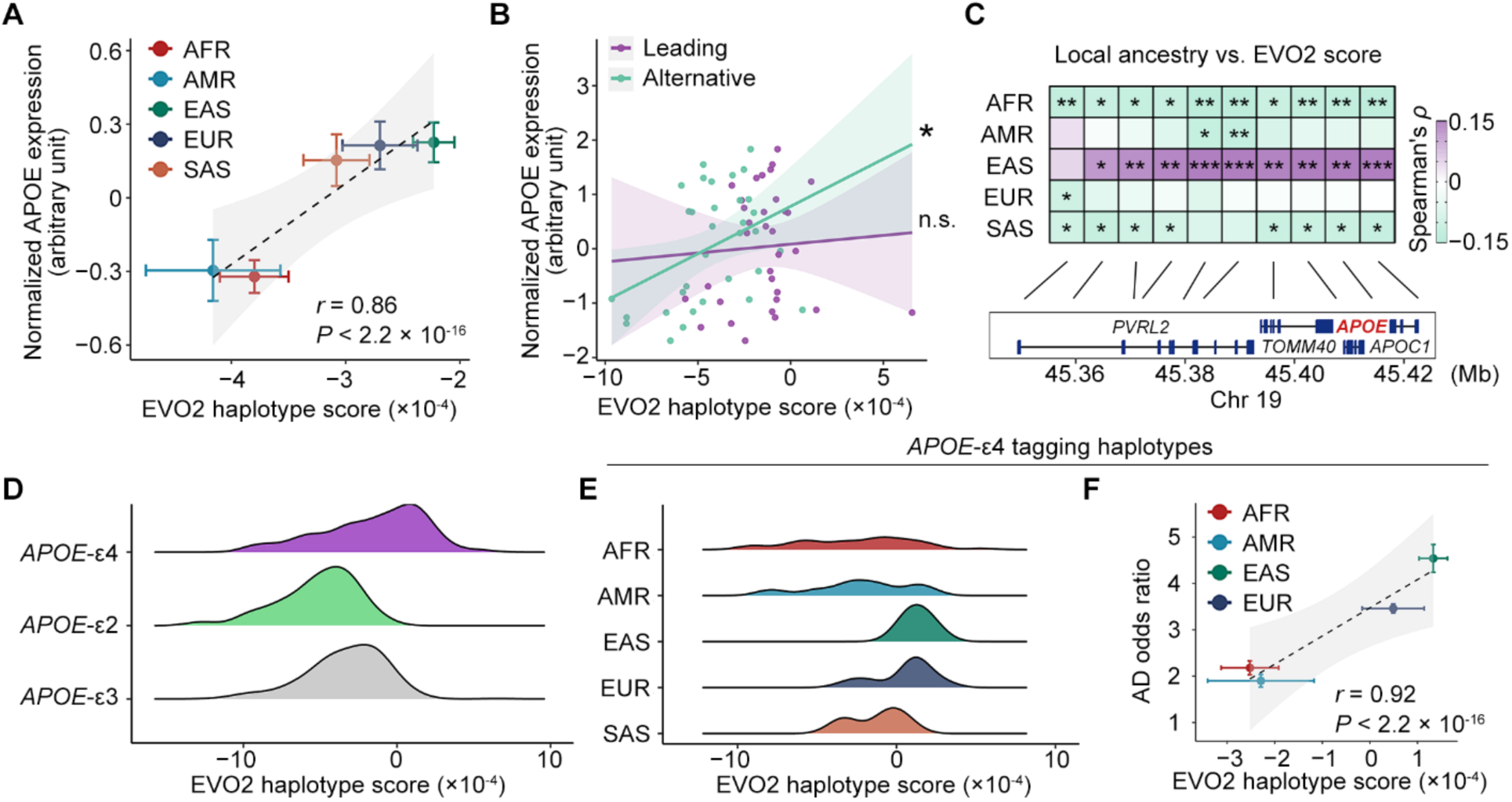
Evo2 Haplotype Scores Across Diverse Ancestry Backgrounds in Pangenome Data. **(A)** Correlation between APOE transcript level and Evo2 haplotype score across five ancestry groups (n = 134, 88, 100, 60, and 72 for AFR, AMR, EAS, EUR, and SAS, respectively). Values and 95 % credible intervals are derived from a Bayesian error-in-variables linear model; Pearson *r* is reported. **(B)** Correlation between APOE transcript level and Evo2 haplotype score in individuals (n = 33) with both expression and haplotype data, assessed with a linear mixed model (\**p* < 0.05). **(C)** Partial correlation between local ancestry and Evo2 haplotype score, tested with Spearman’s rank correlation (458 haplotypes; \**p* < 0.05; \*\**p* < 0.01; \*\*\**p* < 0.001). **(D)** Distribution of Evo2 haplotype scores stratified by *APOE* genotype (n = 357, 39, and 64 for ε2, ε3, and ε4-harboring haplotypes, respectively). **(E)** Distribution of Evo2 scores for the *APOE*-ε4– tagging haplotype, stratified by ancestry (n = 35, 8, 6, 8, and 6 for AFR, AMR, EAS, EUR, and SAS, respectively). **(F)** Correlation between the Alzheimer’s disease odds ratio of *APOE*-ε4 and Evo2 haplotype score across four ancestry groups, estimated with a Bayesian error-in-variables linear model (Pearson *r*). Odds ratios were taken from PMID 37930705. AFR, African; AMR, American; EAS, East Asian; EUR, European; SAS, South Asian.

We next examined whether Evo2 haplotype scores capture ancestry-specific variation at this locus. Sliding-window partial-correlation analysis revealed a consistent positive association with local East-Asian ancestry, while local African ancestry showed the strongest negative association (**Fig. 3C**; **Table S5**). These patterns mirror epidemiological findings that the *APOE*-ε4 allele confers differing Alzheimer’s disease risks across ancestries (*18*), indicating that Evo2 haplotype scores may likewise track baseline AD susceptibility in diverse ancestral backgrounds.

Accordingly, we examined the Evo2 haplotype-score distribution by stratifying haplotypes according to the classical *APOE* isoforms (ε2, ε3, and ε4). Interestingly, haplotypes carrying the ancestral ε4 allele cluster around zero and stretch modestly into the right-hand tail (**Figs. 3D**, **S3**), whereas ε3-, and especially ε2-harboring haplotypes are shifted toward the left-hand side of the distribution (**Figs. 3D**, **S3**). Statistical analysis showed that the distributions of Evo2 haplotype scores differed significantly among the three *APOE* alleles (Kruskal–Wallis rank-sum test, P < 0.001). This pattern corroborates our previous observation at the variant level (**Fig. 2**), suggesting that haplotypes with higher Evo2 scores may likewise be linked to higher AD susceptibility.

We then stratified the ε4-harbouring haplotypes by their ancestral backgrounds. A striking ancestry gradient emerges: median scores decline monotonically from East-Asian (1.33E−04) to European (4.90E−05), South-Asian (−1.24E−04), American (−2.29E−04), and African ancestries (−2.52E−04), mirroring their relative AD risk (**Fig. 3E**) (*18*). Further correlation analysis relating the Evo2 scores of ε4 haplotypes to the reported *APOE*-ε4 odds ratios across ancestries corroborated this observation (4.54, 3.46, 1.90, and 2.18 for East Asian, European, American, and African populations, respectively), revealing an almost perfectly linear effect (Pearson r = 0.92, P < 2 × 10⁻¹⁶; **Fig. 3F**; **Table S6**). Thus, these results indicate the potential of a sequence-only metric to quantify haplotype-level effects on disease susceptibility, even within complex and diverse genetic backgrounds.

### Evo2 haplotype scores explain Alzheimer’s disease risk

We next sought to investigate if the Evo2 score for each specific haplotype reflects the relative AD risk that it confers. Accordingly, we calculated Evo2 haplotype scores for 1,998 individuals from the ADNI cohort, including 722 participants with AD, 604 with mild cognitive impairment (MCI), and 672 cognitively normal controls (CN). Since the majority of *APOE*-ε4 carriers in the general population are heterozygous, we designated, for each individual, the haplotype with the higher Evo2 score as the “leading haplotype,” which may serve as the primary disease-modifying factor. The other haplotype was designated as the “alternative haplotype,” potentially capturing local ancestry effects that modulate the baseline risk associated with this locus. The violin plots revealed that individuals with AD tend to exhibit higher Evo2 haplotype scores compared to those in the CN or MCI groups, suggesting a potential association between the modeled Evo2 haplotype score and disease phenotype (**Fig. 4A**).

**Figure 4.**
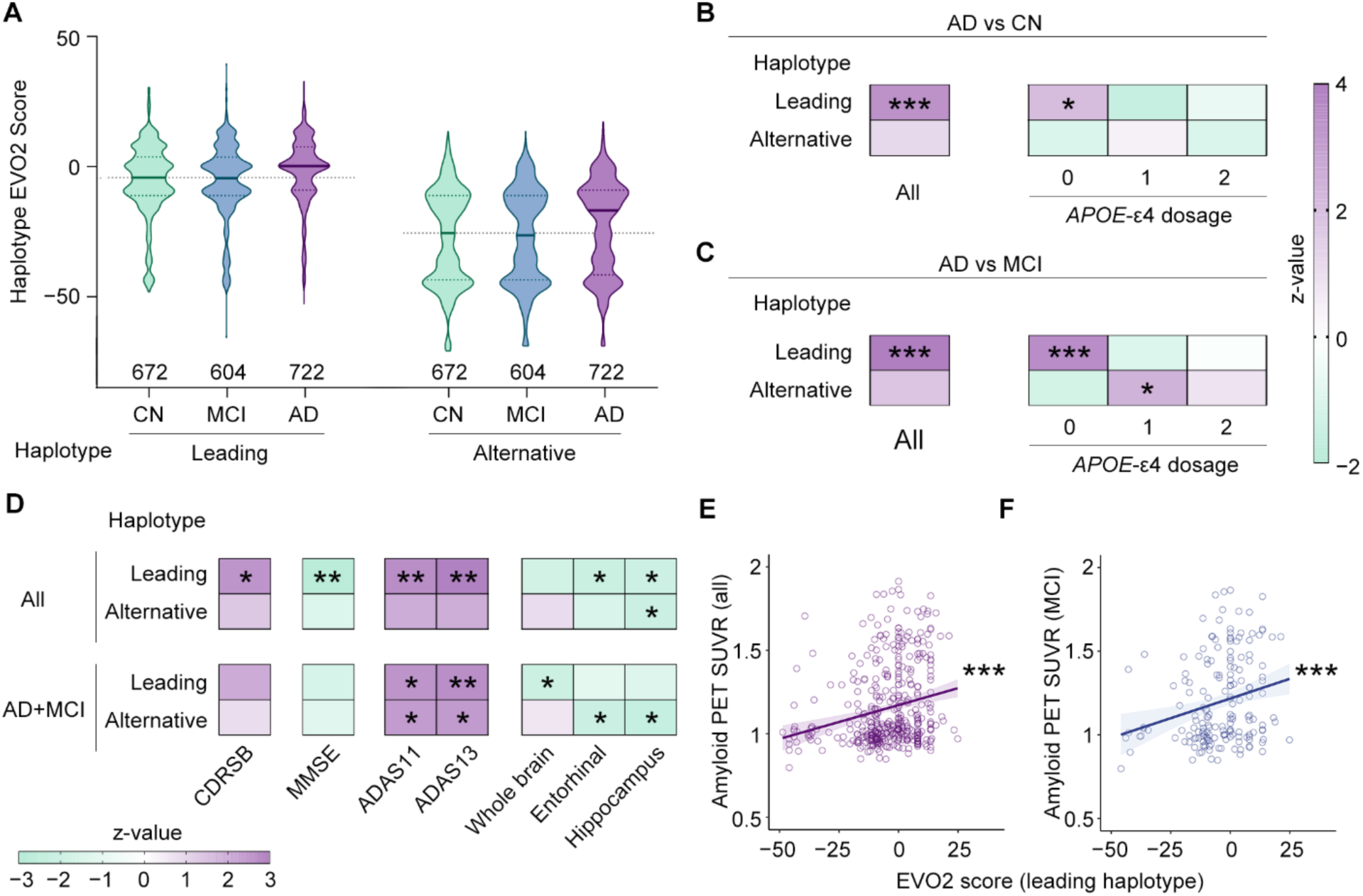
Evo2 Haplotype Scores in Explaining Disease Risk and Endophenotypic Variation in Alzheimer’s Disease Neuroimaging Initiative Data. **(A)** Distribution of Evo2 haplotype scores (leading and alternative haplotypes) across stratified clinical phenotype groups in all individuals. **(B-C)** Association between Evo2 haplotype scores and Alzheimer’s disease (AD; n = 722) in comparison to **(B)** cognitively normal (CN) controls (n = 672), and **(C)** individuals with mild cognitive impairment (MCI; n = 604). Analyses were performed in all participants and stratified by *APOE*-ε4 genotype dosage. Logistic regression; ***p < 0.001, *p < 0.05. **(D)** Association between Evo2 haplotype scores and AD-related endophenotypes. Linear regression; **p < 0.01, *p < 0.05. **(E-F)** Association between Evo2 scores of the leading haplotype and whole-brain amyloid burden at baseline in **(E)** all participants (n = 429) and **(F)** participants with MCI (early and late MCI combined; n = 175). Linear regression; **p < 0.01, *p < 0.05. AD, Alzheimer’s disease; CN, cognitively normal controls; MCI, mild cognitive impairment; CDRSB, Clinical Dementia Rating Scale – Sum of Boxes; MMSE, Mini-Mental State Examination; ADAS, Alzheimer’s Disease Assessment Scale; PET, positron emission tomography; SUVR, standardized uptake value ratio.

To further test this association, we performed a logistic regression analysis. The results confirmed that, in both AD vs. cognitively normal (CN) and AD vs. mild cognitive impairment (MCI) comparisons, the leading Evo2 haplotype score was significantly associated with AD status (P < 0.001; **Figs. 4B–C**; **Table S7**). Notably, stratified analysis based on *APOE*-ε4 allele burden revealed a significant association between Evo2 haplotype scores and AD phenotype among individuals who do not carry *APOE*-ε4 alleles (**Figs. 4B–C**; **Table S7**). This finding reinforces our previous report on the contribution of noncoding haplotypes to AD risk. Together, the results suggest that haplotype effects in the *APOE* region and its surrounding genomic context, as modeled by Evo2, are key to explain individual susceptibility to AD.

We further extended our analysis to additional AD-associated endophenotypes and observed significant associations between haplotype scores and measures of clinical cognitive performance measures, including Clinical Dementia Rating Sum of Boxes (CDR-SB), Mini-Mental State Examination (MMSE), and Alzheimer’s Disease Assessment Scale–Cognitive Subscales 11 and 13 (ADAS-Cog 11, ADAS-Cog 13). Meanwhile, we observed significant association between Evo2 haplotype scores and volumetric changes in brain regions measured by MRI, including whole brain, entorhinal cortex, and hippocampal volumes. These associations were evident in the overall cohort and among individuals with cognitive impairment (AD and MCI; **Fig. 4D**; **Table S8**). Importantly, we also found that the Evo2 score of the leading haplotype strongly predicted individual baseline amyloid burden, as measured by PET imaging, in both the entire cohort (P < 0.001; **Fig. 4E**; **Table S9**) and in individuals with cognitive impairment (P < 0.01; **Fig. 4F**; **Table S9**). Together, these findings suggest that haplotype scores modeled by Evo2 may indicate not only disease onset but also the progression and severity of AD in a cohort that has undergone rigorous clinical assessment.

### Analysis of Evo2 Haplotype Scores in UK Biobank

Next, we examined Evo2’s utility in real-world, population-scale data. We calculated Evo2 haplotype scores for haplotypes defined by 211 AD-associated variants onto 331,925 UK Biobank (UKB) participants who have WGS data and linked electronic health records. Subsets of this cohort also had detailed clinical assessments, brain-imaging metrics, and plasma proteomic profiles.

We first replicated the association between the Evo2 haplotype score and AD in UK Biobank (P < 1 × 10⁻⁸; 2,572 AD cases vs. 117,330 non-demented controls; **Table S10**). Follow-up analyses revealed a moderate association between the score and grey-matter volume in select regions (P < 0.05; n = 29,783; **Figs. 5A**, **S4A**; **Table S11**). The score was likewise also associated with microstructural alterations in AD-vulnerable areas, including the thalamus and amygdala, detected by susceptibility-weighted imaging that captures iron deposition and microvascular stress (P < 0.05; n = 27,749; **Figs. 5A**, **S4B-C**; **Table S12**). Overall, these findings underscore Evo2’s ability to flag subtle neuroanatomical changes that likely precede functional and pathogenic progression.

**Figure 5.**
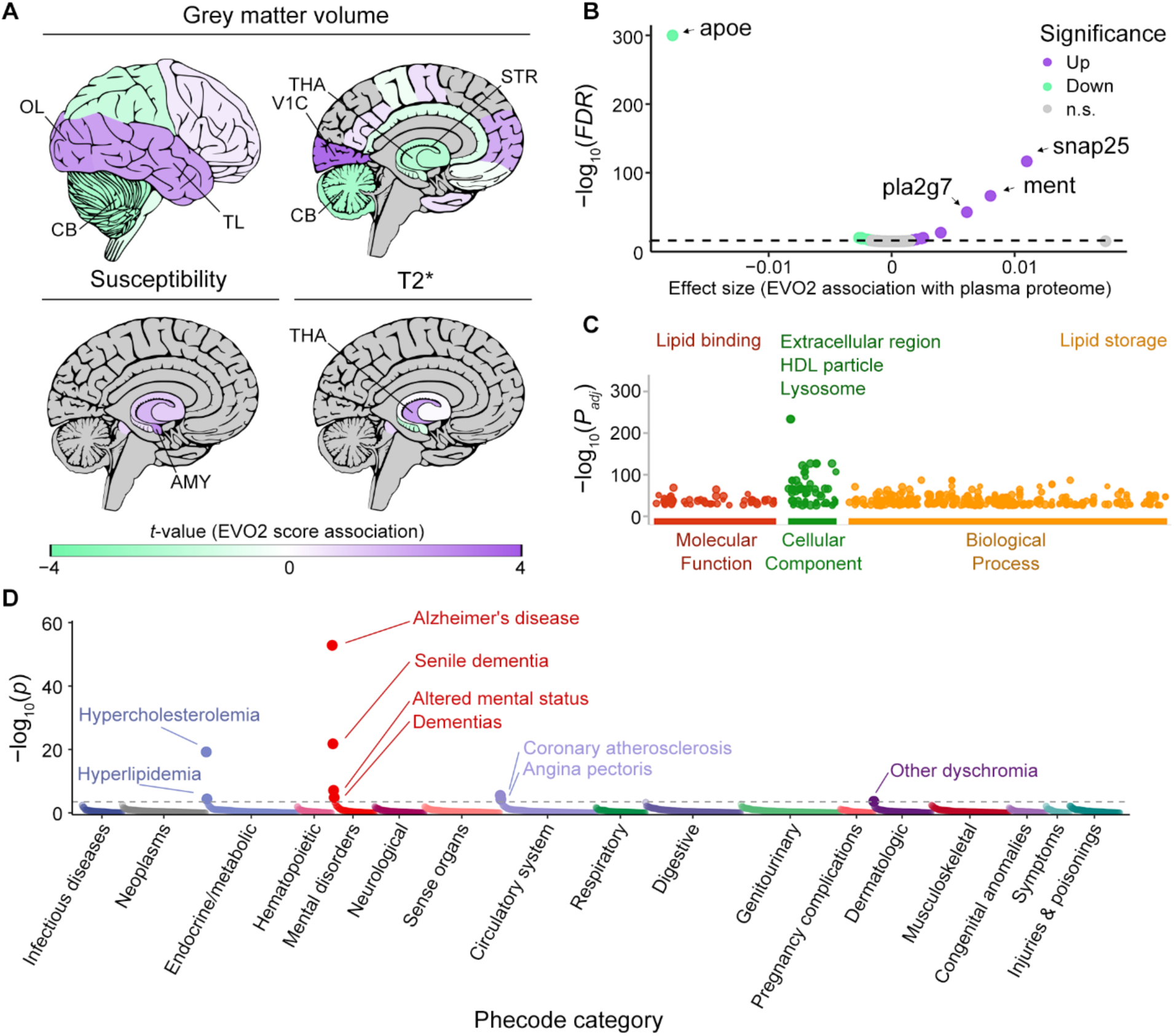
Contribution of Evo2 Haplotype Scores to Phenotypic Variation in UK Biobank Data. **(A)** Associations between the Evo2 haplotype score and brain MRI phenotypes. Upper: regional grey-matter volume from T1-weighted structural scans (n = 29,783). Lower: susceptibility-weighted imaging metrics, including magnetic susceptibility (left) and T2* relaxation time (right) (n = 27,749). Robust linear regression was used; regions with significant effects (p < 0.05) are labelled in black, non-significant regions in grey, and the diverging purple-to-green colour scale denotes positive and negative associations, respectively. **(B)** Volcano plot summarising robust linear-regression results for the association of the Evo2 score (leading haplotype) with plasma protein abundance measured by Olink (2,914 proteins, n = 36,021). **(C)** Gene-ontology enrichment (g:Profiler) for the 30 proteins showing FDR < 0.05 in panel c. **(D)** Phenome-wide association study of the Evo2 score (leading haplotype) across 1,547 PheCodes in 331,925 participants, modelled with logistic regression.

To assess how Evo2 haplotype scores capture inter-individual biological variation at the molecular-pathway level, we correlated them with the abundances of plasma proteins profiled by high-throughput Olink assays (*27*). Of the 2,914 proteins measured in 36,021 participants, 33 are significantly associated with Evo2 scores after FDR correction (q < 0.05), with 28 of them attributable to the leading haplotype (**Fig. 5B**; **Table S13**). For the leading haplotype, the association analysis revealed that the strongest signal came from APOE itself, where higher Evo2 scores were associated with lower plasma ApoE levels. Several other proteins encoded in the locus showed positive correlations, mapping to diverse pathways, such as synaptic function (SNAP25; p = 1.88E-120), protease inhibition (MENT; p = 4.09E-70), and immune-cell metabolism (PLA2G7; p = 3.67E-46). Gene-ontology enrichment analysis echoed these themes, highlighting lipid, cholesterol, and lysosomal processes (**Fig. 5C**; **Table S14**). Collectively, these findings indicate that the modeled Evo2 score reflects baseline activity of specific biological systems potentially underlying the disease’s etiology.

We next studied how the modeled Evo2 haplotype score relates to health outcomes. In a phenome-wide association study of 1,547 PheCodes among 331,925 UK Biobank participants, the Evo2 score showed its strongest links to Alzheimer’s disease, senile dementia, and related cognitive disorders (**Figs. 5D**, **S5**). We also observed weaker yet significant associations with hypercholesterolaemia, coronary atherosclerosis, and dyslipidaemias (**Figs. 5D**). These findings indicate that the Evo2 score quantifies individual susceptibility across a broad range of neurodegenerative, lipid-metabolic, and cardiovascular phenotypes.

## Discussion

In this study, we introduce a first analytical framework that links sequence-based DNA foundation models with population genomics. Leveraging Evo2, we estimate the functional impact of single variants and multi-variant haplotypes across the *APOE* locus and its surrounding genomic landscape. We then project these scores onto a spectrum of resources, including AD GWAS data, Human Pangenome assemblies, deeply phenotyped AD cohort (ADNI), and the population-scale UK Biobank, allowing us to interrogate genetic variation from general populations to disease-specific settings. Our results show that sequence-based models excel at multiple tasks: (i) prioritize variants according to their likely disease relevance; (ii) quantify how haplotypes modulate gene expression at both transcript and protein levels; (iii) predict overall disease susceptibility; and (iv) capture variation in key disease-associated endophenotypes.

Genetics shapes susceptibility to many disorders, yet current genetic analytical tools often fail to capture every causal factor or to quantify precisely how much each locus contributes to disease risk. Most existing methods, and the statistical frameworks behind them, evaluate variant types in isolation, testing their effects separately (*28*, *29*). Such models overlook the nonlinear interactions among variants from the same or different types within a locus, failing to capture the biological reality, that variants act in concert on haplotypes to influence human traits (*30*, *31*, *13*, *32*). Our successful use of sequence-based haplotype scoring offers a powerful alternative: by modeling the combined effects of all variant types within a locus, it shifts association studies from single-variant tests to sequence-level analyses. This strategy not only captures polygenic interactions but also provides a robust way to quantify the impact of rare variations that conventional models lack the power to detect.

Most genomic research has been conducted in populations of European descent with limited representation of other populations (*33*, *34*). Although the current Evo2 model provides a relatively comprehensive genomic atlas across domains of life, it does not adequately capture the diverse genomic landscape across different ancestry backgrounds, as it relies on a single reference genome (GRCh38) to represent the humans (*6*). In recent years, tremendous efforts have been devoted to investigating disease genomics in non-European populations. These joint efforts have enabled a deeper understanding of human biology as well as validated and expanded our findings in non-European populations (*10*, *24*, *35–38*). Using the zero-shot analysis mode, our results demonstrate how sequence-based approaches can be applied to complex genomic backgrounds: Evo2-derived haplotype scores at the *APOE* locus well explained variation in APOE transcript levels as well as the differential impact of the *APOE*-ε4 allele on AD risk across ancestries (**Fig. 3**). We anticipate that further optimization of such sequence-based approaches will greatly enhance their value, providing a promising framework for both transethnic analyses and quantitative studies in admixed populations (*39*).

Many human diseases emerge from a fundamental mismatch between genomic changes accumulated over evolutionary time and our rapid adaptation to modern environments (*40*, *41*). Recent technological breakthroughs now allow us to reshape those environments and, through genome engineering, even potentially altering the (post-natal/somatic) genomes of living organisms (including our own) (*42*, *43*). Evo 2, a large-scale DNA language model trained on genomes spanning the entire tree of life, distills evolutionary signatures into variant-level scores that accurately predict the functional impact of genetic variants across a broad spectrum of biological features. Remarkably, our results show that Evo2 can prioritize AD causal and protective variants by interpreting evolutionary signals encoded across diverse species, without relying on traditional case–control studies (**Fig. 2**). Trained on these cross-species patterns rather than population allele frequencies, Evo2 stands outside the GWAS framework yet offers a complementary layer of evidence that can confirm also extend GWAS discoveries. This finding somehow also raises an intriguing prospect, that is, by interpreting our current genomes, we may forecast future disease susceptibilities and develop proactive precautions, moving beyond the reactive interventions (*44*, *45*, *40*, *6*, *46*).

## Data Availability

Summary statistics from the Alzheimer’s disease GWAS by Jansen et al. are available at https://cncr.nl/research/summary_statistics/;

UK Biobank resources at https://www.ukbiobank.ac.uk/;

Alzheimer’s Disease Neuroimaging Initiative (ADNI) data at https://adni.loni.usc.edu/;

Human Pangenome Reference Consortium datasets at https://humanpangenome.org/;

1000genome expression array dataset at: http://www.ebi.ac.uk/arrayexpress/experiments/E-MTAB-198 and http://www.ebi.ac.uk/arrayexpress/experiments/E-MTAB-264;

Peruvian Genome Project at https://www.ega-archive.org/datasets/EGAD00001007082;

PheWAS Resources repository at https://phewascatalog.org/

## Code Availability

All code used in this study will be available at https://github.com/huthvincent/DNA_LLM_x_population_genomics throughout peer review and following acceptance.

## Supplementary Information

The **Supplementary Information file** contained **Materials and Methods**, **Supplementary Text, 14 Supplementary Tables** and **5 Supplementary Figures**.

## Supporting information

All Supplemental Results

## Acknowledgements

This project was supported in part by the Center for Admixture Science and Technology (NHLBI RM1HG011558). We thank the authors of the Evo2 for their generous guidance and assistance. We are also grateful for the computational resources provided by the Yale Wu Tsai Institute, Yale Center for Research Computing and Yale Biomedical Informatics & Computing.

For Human Pangenome Reference Consortium data, we would like to acknowledge the National Genome Research Institute (NHGRI) for funding the following grants supporting the creation of the human pangenome reference: U41HG010972, U01HG010971, U01HG013760, U01HG013755, U01HG013748, U01HG013744, R01HG011274, and the Human Pangenome Reference Consortium (BioProject ID: PRJNA730823).

For ADNI, data collection and sharing for this project was funded by the Alzheimer’s Disease Neuroimaging Initiative (ADNI) (National Institutes of Health Grant U01 AG024904) and DOD ADNI (Department of Defense award number W81XWH-12-2-0012). ADNI is funded by the National Institute on Aging, the National Institute of Biomedical Imaging and Bioengineering, and through generous contributions from the following: AbbVie, Alzheimer’s Association; Alzheimer’s Drug Discovery Foundation; Araclon Biotech; BioClinica, Inc.; Biogen; Bristol-Myers Squibb Company; CereSpir, Inc.; Cogstate; Eisai Inc.; Elan Pharmaceuticals, Inc.; Eli Lilly and Company; EuroImmun; F. Hoffmann-La Roche Ltd and its affiliated company Genentech, Inc.; Fujirebio; GE Healthcare; IXICO Ltd.; Janssen Alzheimer Immunotherapy Research & Development, LLC.; Johnson & Johnson Pharmaceutical Research & Development LLC.; Lumosity; Lundbeck; Merck & Co., Inc.; Meso Scale Diagnostics, LLC.; NeuroRx Research; Neurotrack Technologies; Novartis Pharmaceuticals Corporation; Pfizer Inc.; Piramal Imaging; Servier; Takeda Pharmaceutical Company; and Transition Therapeutics. The Canadian Institutes of Health Research is providing funds to support ADNI clinical sites in Canada. Private sector contributions are facilitated by the Foundation for the National Institutes of Health (www.fnih.org). The grantee organization is the Northern California Institute for Research and Education, and the study is coordinated by the Alzheimer’s Therapeutic Research Institute at the University of Southern California. ADNI data are disseminated by the Laboratory for Neuro Imaging at the University of Southern California.

This study makes use of data generated by the Peruvian Genome Project from the Peruvian National Institute of Health. A list of authors who contribute to generate the data will be proportionate by request.

We thank Brian L. Hie and Garyk Brixi (Arc Institute and Stanford University) for helpful discussions and insightful comments that improved this manuscript.

X.Z., was supported by CIHR Fellowship (510508). M.M.A. was supported by the SickKids Restracomp Fellowship. S.W.S. holds the Northbridge Chair in Pediatric Research at The Hospital for Sick Children and the University of Toronto.

## Author contributions

Conceptualization: R.Z., X.Z.; Methodology: R.Z., X.Z.; Software: R.Z., X.Z.; Data curation: R.Z., X.Z., H.Z., M.M.A., W.E.; Formal analysis: R.Z., X.Z., M.M.A., W.E.; Investigation: R.Z., X.Z.; Validation: R.Z., X.Z., M.M.A., W.E.; Visualization: R.Z., X.Z., M.M.A., W.E.; Writing – original draft: R.Z., X.Z.; Writing – review & editing: R.Z., X.Z., H.C., H.T., S.W.S., L.O.-M., M.M.A., W.E., J.H.; Resources: ADNI, H.T., S.W.S., L.O.-M.; Supervision: S.W.S., L.O.-M.

