## Supplementary material for "Advancing Human Population Genomics with DNA Foundation Models": All Supplemental Results

**The PDF file includes:**

Materials and Methods
Supplementary Text
Figs. S1 to S5
Tables S1 to S14
References

### Table of Contents


A. Materials and Methods ---- 5

**Evo2 Scoring** ---- 5

*Overview and Zero -Shot Framework* ---- 5

*Notation and Definitions* ---- 5

*Evo2 Variant score* ---- 6

*Evo2 Haplotype (Multiple Variants) Scoring* ---- 7

*Implementation Details* ---- 9

**Study Data and Cohort Description** ---- 11

*Alzheimer's Disease Genome-wide Association Data* ---- 11

*Human Pangenome Reference Consortium* ---- 11

*Lymphoblastoid Cell Line Array Expression Data* ---- 12

*Alzheimer's Disease Neuroimaging Initiative cohort* ---- 12

*UK Biobank Cohort* ---- 12

*Other Reference dataset used in the analysis* ---- 13

**Alzheimer's Disease Genome-Wide Association Data Analysis** ---- 14

**Human Pangenome Project Data Analysis** ---- 15

*Haplotype Extraction and Variant Calling from Human Pangenome Assemblies* -- 15

*Local Ancestry Inference in Human Pangenome Assembly Variant Call* ---- 16

**Alzheimer's Disease Neuroimaging Initiative Cohort Data Analysis** ---- 17

*Evo2 Haplotype Score in Alzheimer's Disease Neuroimaging Initiative Cohort* ---- 18

*Phenotype and Endophenotype Association Analysis in ADNI data* ---- 18

*Evo2 Haplotype Score in UK Biobank Cohort* ---- 19

*Association Between Evo2 Haplotype Scores and AD Phenotype* ---- 20

*Association Between Evo2 Haplotype Scores and Brain MRI Metrics* ---- 20

*Association Between Evo2 Haplotype Scores and Plasma Proteome Data* ---- 21

*Gene Ontology Analysis of Biological Processes Associated with Evo2 Scores* ---- 21

*Phenome-Wide Association Study of Evo2 Haplotype Scores* ---- 21

B. Supplementary Text ---------- 23

*Abbreviations* ---------- 23

C. Supplementary Figure ---------- 25

*Figure S1. Workflow of the Evo2 scoring process* ---------- 25

*Figure S2. Schematic of Evo2 zero-shot scoring* ---------- 26

*Figure S3. Distribution of Evo2 scores for the APOE-ε3 and ε2–tagging haplotype stratified by ancestry in Pangenome dataset* ---------- 27

*Figure S4. Association of Evo2 haplotype score with brain MRI data in the UK Biobank data.* ---------- 28

*Figure S5. Phenome-wide association study results for the Evo2 haplotype score in the UK Biobank data (alternative haplotypes).* ---------- 29

D. Supplementary Table ---------- 30

*Table S1. Cohort and datasets.* ---------- **Error! Bookmark not defined.**

*Table S2. Selected 211 AD-associated variants for Evo2 evaluation from AD GWAS data.* ---------- 31

*Table S3. Mean and SE of Evo2 scores and APOE expression levels across the five major ancestry groups from array expression data.* ---------- 36

*Table S4. Association of Evo2 haplotype scores with APOE expression in Pangenome data.* ---------- 37

*Table S5. Correlation of Evo2 haplotype scores with local ancestry burden estimated using RFMix2 in Pangenome data.* ---------- 38

*Table S6. Mean and SE of Evo2 haplotype scores for APOE-ε4 across the five major ancestry groups in Pangenome data.* ---------- 39

*Table S7. Association between Evo2 haplotype score and Alzheimer's disease in the ADNI data.* ---------- 40

*Table S8. Association of Evo2 haplotype scores with Alzheimer's disease–related endophenotypes in the ADNI data.* ---------- 41

*Table S9. Association of Evo2 haplotype score with PET-measured brain amyloid burden in the ADNI data.* ---------- 43

*Table S10. Association of Evo2 haplotype score with Alzheimer's disease in the UK Biobank data.* ---------- 44

*Table S11. Association of Evo2 haplotype score with regional grey matter volumes (FAST) in the UK Biobank data (p-value < 0.05).* ---------- 45

*Table S12. Association of Evo2 haplotype score with susceptibility weighted brain MRI in the UK Biobank data (p-value $< 0.05$).* ---------------------------------------- 47

*Table S13. Association of Evo2 haplotype score with Olink plasma proteome in the UK Biobank data (FDR $< 0.05$).* -------------------------------------------------------- 48

*Table S14. Phenome-wide association study results for the Evo2 haplotype score in the UK Biobank data (FDR $< 0.05$).* --------------------------------------------------- 50

References------------------------------------------------------------------------------------ 51

### A. Materials and Methods

#### Evo2 Scoring

*Overview and Zero-Shot Framework*

We leveraged the Evo2 model (*1*) to estimate the relative impact of DNA variants without any task-specific fine-tuning (i.e., "zero-shot" mode). Evo2 is trained on genomic data from a wide range of organisms, implicitly learning from large-scale evolutionary patterns to estimate the probability of any given sequence. A key assumption is that variants which significantly reduce (or alter) Evo2's predicted likelihood for a given genomic context may be more disruptive to biological function.

Below, we outline each step of our method for clarity. In section 1, "**Notation and Definition**," we define the reference and variant sequences that are fed into Evo2 and explain how we choose the genomic region around each variant. In section 2, "**Evo2 Variant score**", we introduce our key metrics for assessing whether a mutation is likely to be biologically disruptive. In section 3, "**Evo2 Haplotype (Multiple Variants) Scoring**", we extend the same approach to cases where multiple variations co-occur on the same haplotype. Finally, in section 4, "**Implementation Details**", we implement these steps into a practical workflow, including code snippets and their runtime considerations.

*Notation and Definitions*

Let $\boldsymbol{v}$ be a single-nucleotide variant (SNV) or small insertion/deletion (indel) located at genomic position p. Define:

- $\mathbf{R} = \{r_1, r_2, \ldots, r_L\}$ as the **reference** DNA sequence (the "wild-type"), spanning a context window around p. For instance, a window of $\pm 40$kb from position p yields L = 80001 bases if p is in the middle.
- $\mathbf{V} = \{v_1, v_2, \ldots, v_L\}$ as the **variant** DNA sequence identical to **R** except that at position p, the reference allele(s) are replaced by the alternate allele(s) of interest.

Because Evo2 is an autoregressive language model, it assigns a probability to a full sequence **x** by decomposing it into

$$P(\mathbf{x}) = \prod_{i=2}^{L} P(x_i \mid x_1, x_2, \ldots, x_{i-1}).$$

We define a log-likelihood measure:

$$\mathrm{LL}(\mathbf{x}) = \sum_{i=2}^{L} \log P(x_i \mid x_1, \ldots, x_{i-1}).$$

*Evo2 Variant score*

For a given variant v, we compute:

1. $\mathrm{LL}(\mathbf{R})$, the log-likelihood of the reference window.
2. $\mathrm{LL}(\mathbf{V})$, the log-likelihood of the variant window.

We then define the **Evo2 Variant score** as log-likelihood difference:

$$\Delta\mathrm{LL}(v) = \mathrm{LL}(\mathbf{V}) - \mathrm{LL}(\mathbf{R}).$$

A negative $\Delta\mathrm{LL}(v)$ indicates that Evo2 deems **V** less likely than **R**, potentially implying the variant is more disruptive from an evolutionary standpoint (e.g. *APOE*-ε2). Conversely, a positive ΔLL(v) indicates that Evo2 deems **V** more likely than **R**, suggesting the variant is less disruptive from an evolutionary standpoint (e.g., *APOE*-ε4).

In practice, many analyses use the **absolute value** of this difference, $|\ \Delta\mathrm{LL}(v)\ |$, as a magnitude-based measure of how strongly the variant changes the model's likelihood. Large $|\ \Delta\mathrm{LL}(v)\ |$ indicates a significant departure from sequence patterns that Evo2 has learned to be evolutionarily conserved.

*Evo2 Haplotype (Multiple Variants) Scoring*

If multiple variants $\{v_1, \ldots, v_m\}$ co-occur on the same haplotype, we can define a **haplotype sequence H** in which each relevant position is replaced with its variant allele. Let $\mathrm{LL}(\mathbf{H})$ be the log-likelihood of a haplotype in the specified window, and $\mathrm{LL}(\mathbf{R})$ the log-likelihood of the corresponding reference in the window. The haplotype-based Evo2 difference becomes:

$$\Delta\mathrm{LL}(\text{ haplotype }) = \mathrm{LL}(\mathbf{H}) - \mathrm{LL}(\mathbf{R}).$$

Because multiple variants may have non-linear interactions, the haplotype's $\Delta\mathrm{LL}$ is not always the sum of individual ΔLL values. Nevertheless, for interpretability, we typically report an aggregate haplotype-level ΔLL (since all sequences in this study have the same length), or simply use $\Delta\mathrm{LL}(\text{ haplotype })$ as a single metric capturing the combined effect of all variant alternative alleles constitute the haplotype.

Since the majority of *APOE*-ε4 carriers in the general population are heterozygous, we designated, for each individual, the haplotype with the higher Evo2 score as the "leading haplotype," which may serve as the primary disease-modifying factor. The other haplotype was designated as the "alternative haplotype," potentially capturing local ancestry effects that modulate the baseline risk associated with this locus.

In practice, Evo2 scores（Both Evo2 Variant score and Evo2 Haplotype Scoring）are reported in two equivalent but differently scaled forms:

1. Raw log-likelihood difference ( $\Delta LL$ ).

This is the quantity defined above—either $\Delta LL(v)$ for a single variant or $\Delta LL(H)$ for a haplotype. When all sequence windows in an analysis have identical length (as is the case for every experiment in this paper except one), the raw score is sufficient because length cancels out when comparisons are made.

2. Length-normalized log-likelihood difference ( $\Delta LL/L$ ).

Here the raw $\Delta LL$ is divided by L, the number of nucleotides in the window being evaluated. Normalization converts the score to a per-base log-likelihood change, enabling fair comparison when windows vary in size (e.g., the variants are short insertions or deletions).

Throughout the manuscript we use the raw $\Delta LL$ by default. The sole exception is in the section "Evo2 scores capture haplotype effects across diverse population backgrounds," where we analyze windows extracted from a pangenome graph whose lengths are not uniform. In that context we report normalized $\Delta LL$ so that each haplotype's score is corrected for window length, ensuring that differences reflect variants' effects rather than trivial size effects.

*Implementation Details*

The complete workflow and its accompanying pseudocode for Evo2 scoring are presented in **Fig. S1** and **S2**, respectively.

Hardware and Environment Setup

All Evo2-based computations were performed on a single high-performance server running **Ubuntu 22.04.5 LTS** (kernel 5.19.0-rc6). The server has **2× AMD EPYC 9124 16-Core Processors** (64 total CPU cores, 128 threads), **376 GiB system memory**, and **1× NVIDIA H100** GPU (80 GB VRAM). We verified available memory (≈340 GiB free whenidle) and GPU status (nvidia-smi) prior to each run to ensure sufficient resources for Evo2's scoring of sequences up to ≈80 kb in length.

To mitigate out-of-memory (OOM) events, we conducted most Evo2 queries in single-batch mode (batch_size=1 in the scoring pipeline) and periodically called torch.cuda.empty_cache(). For the *APOE* region, which spans up to 80,001 bp per sequence, this setup was sufficient to run zero-shot scoring on the single H100 GPU. When handling a higher volume of sequences, we carefully monitored GPU memory via nvidia-smi.

All experiments were carried out in a dedicated Conda environment. We exported the complete dependency specification—including Python, CUDA, all library versions, and their channel sources—using conda env export; this file follows the Evo2 recommended configuration available on GitHub(*2*).

Loading Evo2 and Model Checkpoints

We used a local fork of the Evo2 code (https://github.com/ArcInstitute/evo2) with minimal modifications to the original models.py and scoring.py scripts. The core StripedHyen architecture and the associated tokenization logic (via CharLevelTokenizer) remain unaltered.

- **Model Weights**: We primarily worked with the evo2_7b checkpoint. This checkpoint was downloaded once and stored locally.
- **Initialization**: A single call to Evo2(model_name="evo2_7b") in Python loaded the appropriate config (e.g., config.yaml) and merged any checkpoint shards into a .pt file if needed.

All GPU memory management (splitting the model layers, etc.) was handled automatically by the code. Because we used only a single H100 GPU, we did not need to distribute the model across multiple devices. After successful initialization, the script performed a quick test inference to confirm all shards had merged properly.

### Biological Interpretation and Ranking

Finally, variants or haplotypes were ranked by $|\Delta LL|$. A higher absolute Evo2 score suggests that the mutation is more likely to disrupt well-established sequence patterns, possibly affecting function. For noncoding variants, the model may identify potential regulatory disruptions. We integrate these scores into downstream statistical pipelines (e.g., correlation with GWAS effect sizes or disease classification models) to highlight variants or haplotypes of greatest biological concern.

### Study Data and Cohort Description

*Alzheimer's Disease Genome-wide Association Data*

We retrieved the Alzheimer's disease (AD) genome-wide association study (GWAS) results from Jansen et al. (2018) (*3*, *4*). To identify AD-associated variants for evaluating Evo2's credibility in estimating variant and haplotype effects, we retained variants with rsID annotations, a minor allele frequency (MAF) greater than 5%, and located within the *APOE* region (chr19:45,345,000–45,425,000; GRCh37). From this subset, we further selected AD-associated variants by retaining those with a GWAS p-value $< 1\times10^{-4}$ and a false discovery rate (FDR) $< 0.05$, resulting in 211 variants for further investigation (**Table S1**).

*Human Pangenome Reference Consortium*

To assess whether Evo2 capture ancestry-related variation in haplotype structure, we downloaded the Phase 1 and Phase 2 assemblies from the Human Pangenome Reference Consortium (*5*, *6*). The dataset contains 548 haplotype-resolved assemblies (two per individual) representing 274 individuals. By cross-referencing the assembly sample IDs with the International Genome Sample Resource (IGSR) portal (*7*, *8*), we retained 230 individuals for whom super-population labels were available, spanning the five IGSR super-populations.

*Lymphoblastoid Cell Line Array Expression Data*

We retrieved array-based expression profiles for lymphoblastoid cell lines (LCLs) generated by the 1000 Genomes and HapMap projects from IGSR (*9*) and the EMBL-EBI repository (accessions E-MTAB-198 and E-MTAB-264 (*10*, *11*)).

*Alzheimer's Disease Neuroimaging Initiative cohort*

We obtained genotype and phenotype data from the Alzheimer's Disease Neuroimaging Initiative (ADNI) database (*14*) (https://adni.loni.usc.edu/). The ADNI was launched in 2003 as a public-private partnership, led by Principal Investigator Michael W. Weiner, MD. The primary goal of ADNI has been to test whether serial magnetic resonance imaging (MRI), positron emission tomography (PET), other biological markers, and clinical and neuropsychological assessment can be combined to measure the progression of mild cognitive impairment (MCI) and early Alzheimer's disease (AD). For the present study, we included array genotype data obtained from ADNI-1, ADNI-2/GO, and ADNI-3 for analysis. We retained 1,998 individuals, including 722 participants with Alzheimer's disease (AD), 604 with mild cognitive performance (MCI), and 672 cognitively normal controls (CNs), for downstream analysis. The phenotypes of the ADNI participants are from the participants' latest diagnostic records (updated April 2025).

*UK Biobank Cohort*

Genotype, clinical, and multi-omic data were accessed through the UK Biobank Research Analysis Platform on DNAnexus (project ID 41910) (*15*). Phenotypic variables were downloaded with the official Jupyter notebooks provided in the UKB_RAP GitHub repository (https://github.com/dnanexus/UKB_RAP). Specifically, we retrieved the following data from the UKB_RAP to examine the associations between estimated Evo2 haplotype scores, disease status, and related endophenotypic changes: Category 100094: Baseline characteristics; Category 100313: Genotyping process and sample QC; Category 2002: Summary diagnoses; Category 1101: Regional grey-matter volumes (FAST); Category 109: Susceptibility-weighted brain MRI metrics; Category 1839: Protein biomarkers.

For all analyses, we began by applying three quality-control filters recorded under UK Biobank Category 100313: (i) absence of sex-chromosome aneuploidy, (ii) no close kinship with another participant, and (iii) availability of principal-component (PCA) ancestry data. These criteria yielded 333,791 participants for downstream work. Particularly, for the AD association study, cases were defined as individuals carrying an ICD-10 diagnosis of G30 (Alzheimer disease) or F00 (dementia in Alzheimer disease), resulting in 2,572 AD cases. Controls were drawn from the remaining cohort after excluding anyone with ICD-10 codes that could confound cognitive outcomes—namely all dementia codes plus any codes from chapter F (mental and behavioural disorders), chapter G (diseases of the nervous system), chapter C (malignant neoplasms), D00–D49 (in-situ, benign, or uncertain neoplasms), and chapter I (circulatory diseases). The resulting "undemented control" set contains 117,330 participants free of major neurological, psychiatric, neoplastic, and vascular diagnoses.

*Other Reference dataset used in the analysis*

The reference genome sequence for APOE and its surrounding region (chr19:45,345,000–45,425,000; GRCh37) was retrieved using the UCSC "Get DNA in

Window" tool (https://genome.ucsc.edu/cgi-bin/das/hg19/dna?segment=chr19:45345000,45425000) (*16*). The GRCh38 reference was obtained from the Genome Reference Consortium Human Build 38 patch release 14 (GRCh38.p14) (https://ftp.ncbi.nlm.nih.gov/genomes/all/GCA/000/001/405/GCA_000001405.29_GRCh38.p14/GCA_000001405.29_GRCh38.p14_genomic.fna.gz). Super-population and sex annotations for the Pangenome samples were obtained from the IGSR portal (https://www.internationalgenome.org/data-portal/sample). Ancestry-specific odds ratios for the association between *APOE*-ε4 and Alzheimer's disease were extracted from a study (*17*).

### Alzheimer's Disease Genome-Wide Association Data Analysis

For **Fig. 2A**, we calculated Evo2 scores for all 211 Alzheimer's-disease–associated variants (see previous section) and plotted them alongside the original GWAS Z-statistics in a regional plot to compare their spatial patterns of association.

For **Fig. 2B** and **2C**, Pearson correlations were performed in GraphPad Prism (v 8.0.1) to examine the relationship between Evo2 scores and GWAS signals. We correlated (i) the GWAS Z-scores with absolute Evo2 scores to test whether Evo2 prioritizes the most influential variants, and (ii) GWAS effect sizes (β) with Evo2 scores to assess whether Evo2 reflects effect-size magnitude. Analyses were conducted on the full variant set (n = 211, **Fig. 2B**) and on a clumped subset (n = 28, **Fig. 2C**) created with PLINK's clumping procedure (*18*); within each clump, the variant with the highest absolute Evo2 score served as the representative tagging variant.

For **Fig. 2D** and **2E**, functional annotation was performed with SNPnexus (SNPnexus (https://www.snp-nexus.org/v4/; Ensembl build) to assign gene and protein consequences (*19*). Variants in non-coding regions (intronic or intergenic) were further annotated with the ENCODE SCREEN database (https://screen.wenglab.org/) to evaluate overlap with candidate cis-regulatory elements (*20*). Differences in Evo2 scores across functional categories were tested using one-way ANOVA followed by the Benjamini–Krieger–Yekutieli two-stage linear step-up post-hoc procedure in GraphPad Prism.

### Human Pangenome Project Data Analysis

#### *Haplotype Extraction and Variant Calling from Human Pangenome Assemblies*

For 460 haplotype assemblies from 230 individuals, the APOE locus was isolated by remapping the GRCh38 interval chr19:44,841,743–44,921,743 (equivalent to chr19:45,345,000–45,425,000 in GRCh37) to each assembly with minimap2 v2.26 (*-x asm20 -t 20 --secondary=no*) (*21*). From the resulting PAF alignment files, we identified a consensus region (bases 22–79,918 within the selected interval) that is present in every pangenome assembly. Thus, we extracted (i) bases 22–79,918 of the mapped reference segment and (ii) the corresponding assembly segment from the resulting PAF alignment files. The former served as the reference sequence and the latter as the target sequence for subsequent Evo2 scoring. Please refer to the section "**Lymphoblastoid Cell Line Array Expression Data Analysis**" below for the association between pangenome data and expression data shown in **Figs. 3A–B**.

To derive *APOE* genotypes (*APOE*-ε2, *APOE*-ε3, *APOE*-ε4) from each assembly, we realigned every extracted haplotype segment to its corresponding reference with minimap2 v2.26 (-a -x asm20 -t 20 --secondary=no). The resulting BAM files were sorted and indexed with samtools, and variants were then called with bcftools: bcftools mpileup (--no-BAQ) followed by bcftools call (--ploidy 1) (*22*). This workflow produced haploid VCF files containing the APOE coding variants for each assembly. *APOE* isoforms (ε2, ε3, ε4) were assigned according to the allele combination at the two defining SNPs, rs429358 and rs7412. To evaluate whether EVO2 haplotype scores differed among haplotypes harboring different *APOE* isoforms (ε2, ε3, ε4), we ran a Kruskal–Wallis rank-sum test with *kruskal.test()* from the *stats* package, followed by Dunn's pair-wise post-hoc comparisons using *dunn.test()* from the *dunn.test* package.

*Local Ancestry Inference in Human Pangenome Assembly Variant Call*

We ran RFmix2 (*23*) to infer local ancestry within the *APOE* region on chromosome 19. The phased GRCh38 VCF file was converted to BCF format and indexed. The reference panel consisted of ~400 unrelated individuals from each ancestry group: African (n = 425), European (n = 404), East Asian (n = 405), South Asian (n = 402), and American (n = 64) individuals from the 1000 Genomes Project (*24*), we also added for the American ancestry 358 individuals from the Peruvian Genome Project (*25*).

For Fig. 3C, we evaluated the correlation between local ancestry probabilities estimated with RFMix2 (see above section) and Evo2 haplotype scores across all haplotypes in the Pangenome cohort. At each locus, partial correlations were calculated with the *pcor.test()* function from the *ppcor* package, treating the focal ancestry as the variable of interest and using the remaining ancestries as covariates. We reported the Spearman correlation coefficient and its corresponding *p*-value for every ancestry-locus pair.

**Lymphoblastoid Cell Line Array Expression Data Analysis**

Before downstream analyses, the expression intensities were log2-transformed and quantile-normalized with the normalizeBetweenArrays() function in the limma R package (*12*). Matching the expression data to IGSR metadata left 587 individuals, 33 of whom are also included in the Pangenome cohort. For downstream analyses, APOE expression values were extracted from the normalized matrix and rank-inverse-normalized using the RankNorm() function from the RNOmni (*13*) package.

For **Fig. 3A** correlation analysis, we first calculated the mean and standard error of the Evo2 haplotype score and of the rank-normalized APOE transcript level within each of the five ancestry groups. We then assessed their association by fitting a Gaussian Bayesian errors-in-variables model with the *brm()* function in the *brms* package (*26*), explicitly incorporating the standard errors of both variables as measurement error. A two-tailed Bayesian *p*-value was obtained from the posterior distribution of the slope (β) as twice the smaller fraction of draws above or below zero. The strength of the correlation was quantified with a Bayesian Pearson correlation calculated with 6 000 posterior iterations.

For **Fig. 3B** association analysis, we used the 33 participants who had both Evo2 haplotype scores and APOE expression measurements. Separate linear mixed-effects models were fitted for the leading and alternative haplotypes with the lmer() function in the lmerTest package (*27*), treating the Evo2 score as a fixed predictor of APOE expression and sex as a random effect.

### Alzheimer's Disease Neuroimaging Initiative Cohort Data Analysis

*Evo2 Haplotype Score in Alzheimer's Disease Neuroimaging Initiative Cohort*

We downloaded genotype array datasets (WGS_Omni25_BIN_wo_ConsentsIssues, ADNI_cluster_01_forward_757LONI, ADNI_GO_2_Forward_Bin, ADNI_GO2_GWAS_2nd_orig_BIN, and ADNI3_PLINK_Final) and the whole-genome sequencing (WGS) dataset from the ADNI database. For the array data, we first converted genomic coordinates to GRCh38 using *liftOverPlink.py* (https://raw.githubusercontent.com/Shicheng-Guo/GscPythonUtility/master/liftOverPlink.py) (*28*). We then used PLINK 1.9 to extract variants within the APOE region (chr19:44,400,000–45,400,000; GRCh38) with a missing rate below 10%. Using PLINK 2, we aligned alleles to the reference genome with the *--ref-from-fa* option and converted the data to unphased VCF format (*--recode vcf*). The resulting VCFs were further processed with *bcftools +fixref* to correct reference allele mismatches. The corrected array-based VCF files and the WGS dataset were then submitted to the Michigan Imputation Server 2 (https://imputationserver.sph.umich.edu/) for phasing and imputation using the HRC r1.1 reference panel (hg19) (*29*, *30*). For individuals with genotype data from multiple arrays, we retained data from the array with the largest number of variants submitted for imputation. We then extracted the phased genotype information for 211 AD–associated variants in all ADNI participants and generated the haplotype-level Evo2 scores from the phased genotypes. The haplotype with the higher Evo2 score was designated the leading haplotype, and the other was designated the alternative haplotype.

*Phenotype and Endophenotype Association Analysis in ADNI data*

For **Fig. 4B–C**, logistic regression (glm in R) tested the association between Evo2 haplotype scores and diagnosis (AD versus CN in **Fig. 4B** and AD versus MCI in **Fig. 4C**) while adjusting for age, sex, years of education, and ancestry principal components. Analyses were further stratified by *APOE* ε4-allele dosage (0, 1, or 2 copies).

For **Fig. 4D–F**, robust linear regression (lmrob in the robustbase package) related Evo2 haplotype scores to quantitative traits , including Clinical Dementia Rating Sum of Boxes (CDR-SB), Mini-Mental State Examination (MMSE), and Alzheimer's Disease Assessment Scale–Cognitive Subscales 11 and 13 (ADAS-Cog 11, ADAS-Cog 13) (*31*). The same covariates were used as in the diagnostic models. Intracranial volume was added when modeling MRI measures (entorhinal cortex, hippocampal, and whole-brain volumes; **Fig. 4D**). For amyloid-PET SUVR outcomes (UCBERKELEY_AMY_6MM; **Fig. 4E–F**), SUMMARY_VOLUME was included to control region size.

### UK Biobank Cohort Data Analysis

#### *Evo2 Haplotype Score in UK Biobank Cohort*

WGS variants spanning the *APOE* locus and its flanking region (chr19: 44,830,000 – 44,930,000; GRCh38) with minor-allele frequency (MAF) > 1 % in the UK Biobank (UKB) cohort were extracted from GraphTyper population-level pVCF files (Data-Field 23374; n = 490,155; from ukb23374_c19_b[2240-2247]._v1.vcf.gz) with *Swiss-Army-Knife*. The resulting VCFs were concatenated and sorted with *bcftools sort*, then normalized with *bcftools norm* to split multiallelic sites into separate records, using the GRCh38 reference genome. rsIDs were annotated with *bcftools annotate* against dbSNP v165. Genotypes were subsequently phased and refined with *Beagle* (22 Jul 22) using the 1000 Genomes Phase 3 panel and the GRCh38 genetic map (*32*).

From the phased VCF we extracted genotypes for 211 Alzheimer's-disease–associated sites and retained those with *Beagle* dosage $R^2$ (DR2) > 0.6, leaving 203 high-quality variants for Evo2 haplotype scoring. To reduce the computational load, Evo2 scores for UKB were computed in two stages. First, haplotypes defined by the 203 phased variants were enumerated and assigned unique indices. Second, an Evo2 score was calculated for each unique haplotype and then mapped back to individuals via these indices.

*Association Between Evo2 Haplotype Scores and AD Phenotype*

In **Table S10**, association analysis between Evo2 haplotype score (for leading haplotypes and alternative haplotypes) and AD was conducted in 2,572 AD cases and 117,330 non-demented controls using logistic regression implemented by *glm()*, with age, sex, and population structure represented by top-5 principal components as covariates.

*Association Between Evo2 Haplotype Scores and Brain MRI Metrics*

For **Fig. 5A** (upper panel) and **Fig. S2A**, we assessed the association between Evo2 haplotype scores and regional grey-matter volume derived from T1-weighted structural MRI (n = 29,783) using robust linear regression (*lmrob* from *robustabase* package) (*33*). Rank-based inverse-normalized regional grey-matter volume was the outcome, Evo2 haplotype scores for both leading and alternative haplotypes were the predictors. Sex, age, the first five ancestry principal components, and rank-based inverse-normalized total grey-matter volume were included as covariates.

For **Fig. 5A** (lower panel) and **Fig. S2B-C,** we assessed the association between Evo2 haplotype scores and susceptibility-weighted imaging metrics (n = 27,749), including magnetic susceptibility and T2* relaxation time, using robust linear regression (lmrob from *robustabase* package) (*34*). Rank-based inverse-normalized regional data was the outcome, Evo2 haplotype scores for both leading and alternative haplotypes were the predictors. Sex, age, the first five ancestry principal components were included as covariates.

*Association Between Evo2 Haplotype Scores and Plasma Proteome Data*

For **Fig. 5B**, we assessed the association between Evo2 haplotype scores and plasma proteome data measured by Olink platform (2,914 proteins, n = 36,021), using robust linear regression (lmrob) (*35*). Rank-based inverse-normalized protein level data was the outcome, Evo2 haplotype scores for both leading and alternative haplotypes were the predictors. Sex, age, the first five ancestry principal components were included as covariates.

*Gene Ontology Analysis of Biological Processes Associated with Evo2 Scores*

For **Fig. 5C**, we conducted gene-set enrichment analysis with *gprofiler2* on the 28 proteins significantly associated with the leading haplotype (FDR < 0.05) (*36*). Gene Ontology terms from the Biological Process (BP), Molecular Function (MF), and Cellular Component (CC) categories were considered, and only terms achieving FDR < 0.05 are reported.

*Phenome-Wide Association Study of Evo2 Haplotype Scores*

For **Fig. 5D** and **Fig. S5**, we performed a phenome-wide association study (PheWAS) of Evo2 haplotype scores using UK Biobank diagnosis data (Category 1101). All primary and secondary ICD-10 codes were aggregated and mapped to PheCodes with the PheCode Map 1.2 (PheWAS Resources, https://phewascatalog.org/) (*37*). Logistic regression models were then fitted for 1,547 phenotypes in 331,925 participants, treating each PheCode as a binary outcome. Evo2 scores for the leading and alternative haplotypes served as predictors, with sex, age, and the top five ancestry principal components included as covariates.

**Data Visualization**

Regional plots of GWAS signals and Evo2 score distributions (**Fig. 2A**) were generated with the *locuszoom()* function in *topr* package (*38*), using LD information retrieved via *LDproxy()* from *LDlinkR* (*39*). Correlation plots were created with ggplot2 (*40*) (**Fig. 2B–C**, **3A–B**, **3F**) or GraphPad Prism v8.0.1 (**Fig. 4E–F**). Dot plots (**Fig. 2D–E**), heatmaps (**Fig. 3C**, **4B–D**), and violin plots (**Fig. 4A**) were also produced in *GraphPad Prism*. Histograms (**Fig. 3D–E**) and the volcano plot (**Fig. 5B**) were drawn with *ggplot2*. The brain heat map (**Fig. 5A** and **S4**) was generated using the *cerebroViz* (*41*) function *cerebroViz()*. The GO enrichment plot (**Fig. 5C**) was produced with gprofiler2's *gostplot()*. And PheWAS results (**Fig. 5D** and **S5**) were visualized with *ggplot2*.

### B. Supplementary Text

*Abbreviations*

**1. Clinical status & cognitive assessments**

**AD**: Alzheimer's Disease
**MCI**: Mild Cognitive Impairment
**CN**: Cognitively Normal
**ADAS11/ADAS13**: Alzheimer's Disease Assessment Scale (11- & 13-item versions)
**CDRSB**: Clinical Dementia Rating Scale, Sum of Boxes

**2. Research consortia, cohorts & reference resources**

**ADNI**: Alzheimer's Disease Neuroimaging Initiative
**UKB**: UK Biobank
**IGSR**: International Genome Sample Resource
**HRC**: Haplotype Reference Consortium

**3. Population (super-population) codes**

**AFR**: African
**AMR**: American
**EAS**: East Asian
**EUR**: European
**SAS**: South Asian

**4. Imaging modalities & derived metrics**

**MRI**: Magnetic Resonance Imaging
**PET**: Positron Emission Tomography
**SWI**: Susceptibility-Weighted Imaging
**SUVR**: Standardized Uptake Value Ratio

**5. Brain Regions / Segmentations (following cerebroViz)**

**A1C**: primary auditory cortex
**AMY**: amygdala
**ANG**: angular gyrus
**BS**: brainstem
**CAU**: caudate
**CB**: cerebellum
**CNG**: anterior cingulate cortex
**DFC**: dorsolateral prefrontal cortex
**FL**: frontal lobe
**HIP**: hippocampus
**HTH**: hypothalamus
**IPC**: inferior parietal cortex
**ITC**: inferior temporal cortex
**M1C**: primary motor cortex
**MED**: medulla oblongata
**MFC**: medial prefrontal cortex
**OL**: occipital lobe
**OFC**: orbital frontal cortex
**PL**: parietal lobe

**PIT**: pituitary gland
**PUT**: putamen
**PON**: pons
**S1C**: primary somatosensory cortex
**SN**: substantia nigra
**STC**: posterior (caudal) superior temporal cortex
**STR**: striatum
**TL**: temporal lobe
**THA**: thalamus
**V1C**: primary visual cortex
**VFC**: ventrolateral prefrontal cortex

**6. Molecular biology & genomic data types**

**DNA**: Deoxyribonucleic Acid
**WGS**: Whole-Genome Sequencing
**VCF**: Variant Call Format
**SNP** / **SNV**: Single-Nucleotide Polymorphism / Variant
**LCL**: Lymphoblastoid Cell Line
**MHC**: Major Histocompatibility Complex
**bp** / **kbp**: Base Pair / Kilo-Base Pair

**7. Genomic & phenomic study designs / bioinformatic methods**

**GWAS**: Genome-Wide Association Study
**PheWAS**: Phenome-Wide Association Study
**PCA**: Principal Component Analysis
**RFmix**: Random-Forest Mixture (local-ancestry inference)

**8. Functional genomics & regulatory annotations**

**CA**: Chromatin Accessibility
**PLS**: Promoter-Like
**pELS**: Proximal Enhancer-Lik
**dELS**: Distal Enhancer-Like Signatures
**GO**: Gene Ontology

**9. Computing, AI & hardware acceleration**

**AI**: Artificial Intelligence
**LLM**: Large Language Model
**GPU**: Graphics Processing Unit
**CUDA**: Compute Unified Device Architecture

**10. Statistical & frequency measures**

**FDR**: False Discovery Rate
**MAF**: Minor Allele Frequency

**11. Genes / proteins**

**APOE**: Apolipoprotein E

### C. Supplementary Figure

*Figure S1. Workflow of the Evo2 scoring process*

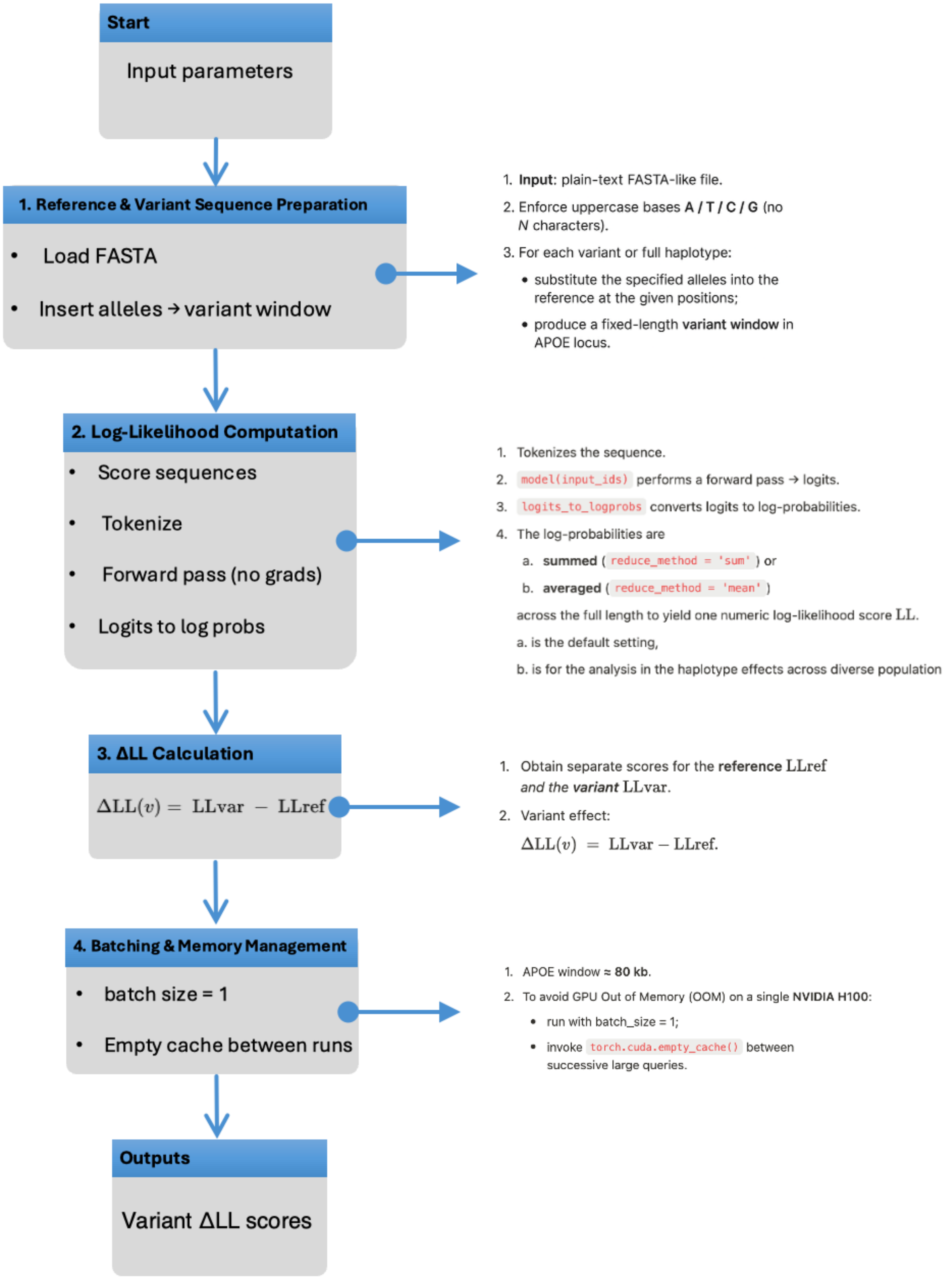

*Figure S2. Schematic of Evo2 zero-shot scoring*

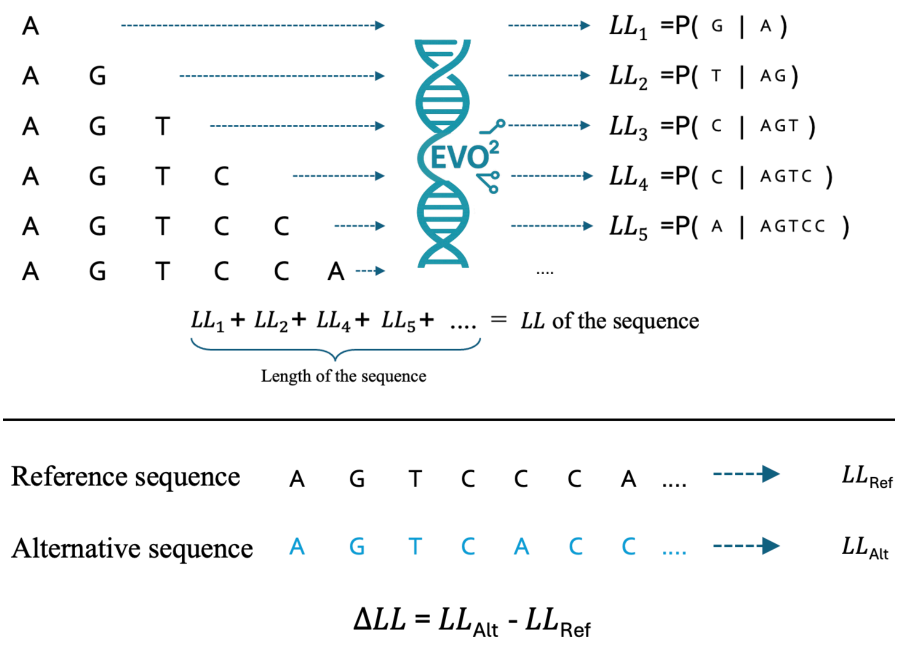


We use Evo2 to estimates the impact of mutations without task-specific training ("zero-shot"). At each position $i$, the model outputs the conditional probability $P(x_i \mid x_{1\ldots i-1})$; taking the natural log yields the site-specific log-likelihood $LL_i$. Summing these values across all positions produces the sequence log-likelihood $LL_{\mathrm{seq}} = \sum_i LL_i$. We independently compute $LL_{Ref}$ for the reference sequence and $LL_{Alt}$ for a mutated sequence, then define the Evo2 score as $\Delta LL = LL_{\mathrm{Alt}} - LL_{\mathrm{Ref}}$.

*Figure S3. Distribution of Evo2 scores for the APOE-ε3 and ε2–tagging haplotype stratified by ancestry in Pangenome dataset*

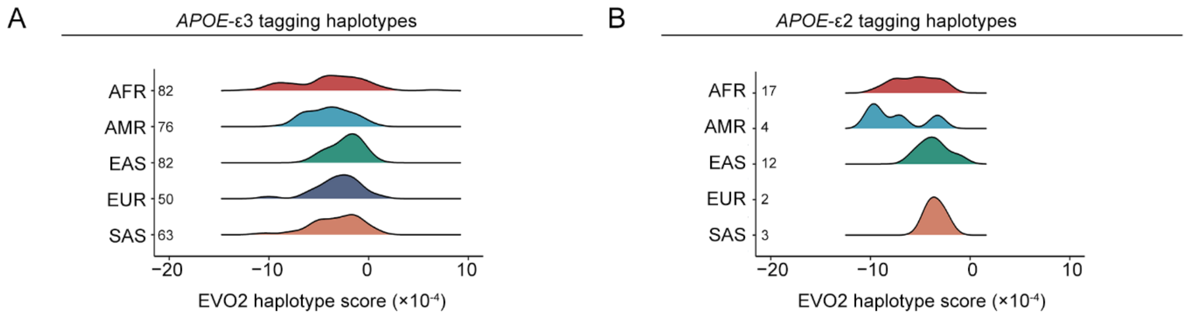


Distribution of Evo2 scores for **(A)** the APOE-ε3–tagging haplotype and **(B)** the APOE-ε2–tagging haplotype, stratified by ancestry. Sample sizes for each ancestry group are indicated on the plots. AFR, African; AMR, American; EAS, East Asian; EUR, European; SAS, South Asian.

*Figure S4. Association of Evo2 haplotype score with brain MRI data in the UK Biobank data.*

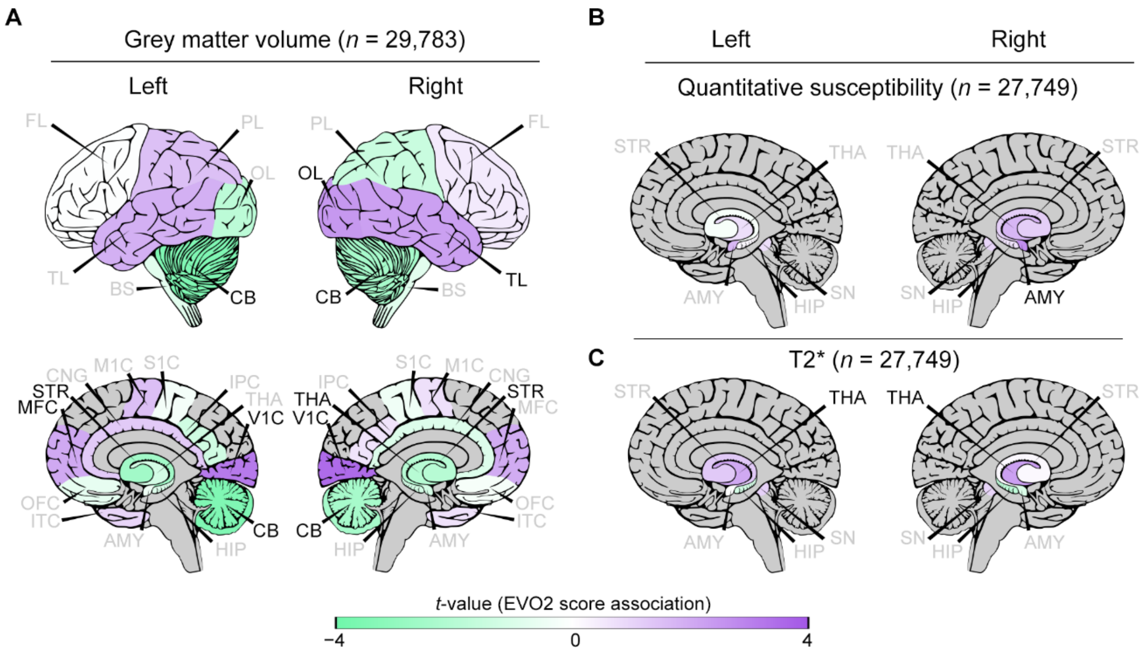


Association between Evo2 haplotype score and brain MRI measures from the UK Biobank. Brain colour map showed results from **(A)** regional grey-matter volume on T1-weighted scans ($n = 29{,}783$) and susceptibility-weighted imaging metrics ($n = 27{,}749$), including **(B)** magnetic susceptibility and **(C)** T2* relaxation time. Panels **A** and **C** show results for the leading haplotype; panel **B** shows results for alternative haplotypes. Robust linear regression was applied. Brain regions with $P < 0.05$ are labelled in black; non-significant regions are in grey. Purple-to-green diverging colour scale represents positive (purple) and negative (green) associations. Region abbreviations follow the scheme in https://ethanbahl.github.io/cerebroViz/. Please refer to the **Abbreviations** section in the **Supplementary Text** for the full names of the corresponding brain regions. MRI, magnetic resonance imaging; n, sample size.

*Figure S5. Phenome-wide association study results for the Evo2 haplotype score in the UK Biobank data (alternative haplotypes).*

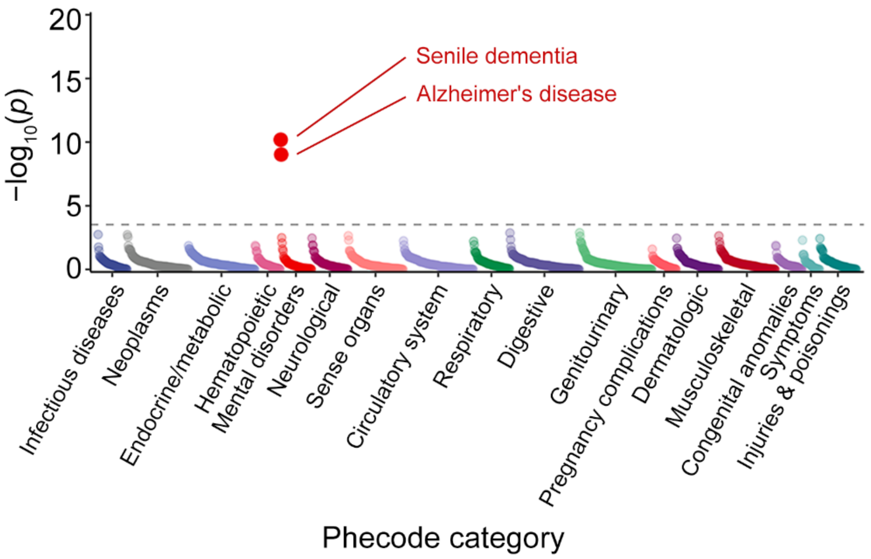


Phenome-wide association study of the Evo2 score for alternative haplotype across 1,547 PheCodes in 331,925 participants, Logistic regression was applied.

### D. Supplementary Table

*Table S1. Cohort and datasets.*

**AD GWAS dataset**

| Name | N (case) | N (control) | | Ref (PMID) |
|---|---|---|---|---|
| AD_sumstats_Jansen etal_2019sept | 71,880 | 383,378 | | 30617256(*4*), |

**Pangenome dataset**

| Name | N (haplotype assembly) | Ref (PMID) |
|---|---|---|
| HPRC | 230 | 37165242(*6*), |

**1000genome expression array dataset**

| Name | N | Ref (PMID) |
|---|---|---|
| LCL_array_expression | 587 | 20220756; 20369022(*42*, *43*), |

**Alzheimer's Disease Neuroimaging Initiative**

| Items | CN | MCI | AD | Ref (PMID) |
|---|---|---|---|---|
| **N** | 672 | 604 | 722 | 20451875(*44*); 29572282(*45*) |
| **Age (SD)** | 77.0 (7.3) | 77.7 (8.1) | 77.8 (7.8) | |
| **Female (%)** | 374 (55.7%) | 247 (40.9%) | 305 (42.2%) | |
| **Education Year (SD)** | 16.7 (2.5) | 15.9 (2.9) | 15.6 (2.9) | |
| ***APOE*-ε4 carrier (%)** | 195 (29.0%) | 241 (39.9%) | 462 (64.0%) | |
| ***APOE*-ε2 carrier (%)** | 88 (13.1%) | 56 (9.3%) | 43 (6.0%) | |

**UK Biobank data**

| Items | N | Ref |
|---|---|---|
| Unrelated participants passed QC | 333,791 | Category 100313 |
| Regional grey matter volumes (FAST) | 29,783 | Category 1101 |
| Susceptibility weighted brain MRI | 27,749 | Category 109 |
| Protein biomarkers | 36,021 | Category 1839 |
| Summary Diagnoses | 331,925 | Category 2002 |

AD, Alzheimer’s disease; CN, cognitively normal; HPRC, Human Pangenome Reference Consortium; LCL, lymphoblastoid cell line; MCI, mild cognitive impairment; MRI, magnetic resonance imaging; N, sample size; PMID, PubMed ID; QC, quality control; Ref, reference; SD, standard deviation.

*Table S2. Selected 211 AD-associated variants for Evo2 evaluation from AD GWAS data.*

| *Index* | *rsID* | *Chr* | *BP (GRCh37)* | *A1* | *A2* | *Evo2_score* |
|---|---|---|---|---|---|---|
| *1* | rs2199575 | 19 | 45345623 | A | G | -0.1641 |
| *2* | rs2199576 | 19 | 45345708 | G | A | -0.1331 |
| *3* | rs2376518 | 19 | 45345773 | T | C | 1.4619 |
| *4* | rs111371860 | 19 | 45345787 | T | A | 1.4219 |
| *5* | rs1985096 | 19 | 45346551 | A | T | 7.8419 |
| *6* | rs1001611 | 19 | 45346768 | A | G | 1.8419 |
| *7* | rs11668738 | 19 | 45347561 | T | G | 4.8819 |
| *8* | rs41289510 | 19 | 45347636 | A | G | -0.8831 |
| *9* | rs4452060 | 19 | 45347911 | A | C | 4.0859 |
| *10* | rs10426423 | 19 | 45348253 | T | C | -2.0181 |
| *11* | rs12162222 | 19 | 45348522 | T | G | -0.4181 |
| *12* | rs77241309 | 19 | 45349177 | C | G | -1.0781 |
| *13* | rs2927472 | 19 | 45349369 | T | C | 8.0619 |
| *14* | rs3810143 | 19 | 45349402 | C | T | 3.0019 |
| *15* | rs2306149 | 19 | 45349963 | A | C | 1.9819 |
| *16* | rs1871047 | 19 | 45351746 | G | A | 7.2319 |
| *17* | rs2972569 | 19 | 45351891 | A | G | 2.3619 |
| *18* | rs1871046 | 19 | 45351937 | C | T | 4.6879 |
| *19* | rs7255063 | 19 | 45352419 | C | G | 3.5719 |
| *20* | rs12974942 | 19 | 45352487 | T | A | 5.7969 |
| *21* | rs4802240 | 19 | 45352804 | T | C | 4.4119 |
| *22* | rs1531516 | 19 | 45353261 | G | C | 2.5319 |
| *23* | rs57537848 | 19 | 45354044 | T | G | 0.8319 |
| *24* | rs2972566 | 19 | 45354238 | G | C | 0.9119 |
| *25* | rs11666329 | 19 | 45354296 | A | G | 2.2319 |
| *26* | rs2927469 | 19 | 45355552 | A | G | 2.4769 |
| *27* | rs11667610 | 19 | 45355595 | T | C | 8.4059 |
| *28* | rs2972559 | 19 | 45355721 | G | C | 0.7019 |
| *29* | rs4802241 | 19 | 45355743 | C | A | -7.0581 |
| *30* | rs2972558 | 19 | 45356141 | C | T | -0.3981 |
| *31* | rs73050205 | 19 | 45356464 | A | T | -6.5231 |
| *32* | rs10421035 | 19 | 45356674 | G | A | 5.3419 |
| *33* | rs35396326 | 19 | 45357003 | G | C | -2.2081 |
| *34* | rs4803763 | 19 | 45357291 | C | G | 5.5219 |
| *35* | rs4803764 | 19 | 45357377 | C | T | -2.8681 |
| *36* | rs2927468 | 19 | 45357939 | A | G | 5.9059 |
| *37* | rs7249933 | 19 | 45358235 | T | C | -1.1281 |
| *38* | rs10410835 | 19 | 45358353 | T | C | 7.1219 |
| *39* | rs12981508 | 19 | 45358500 | A | G | 1.7519 |
| *40* | rs56317818 | 19 | 45359586 | T | C | 3.0719 |
| *41* | rs12462573 | 19 | 45359706 | A | G | 6.2519 |
| *42* | rs2972557 | 19 | 45360573 | G | A | 8.8359 |
| *43* | rs8112526 | 19 | 45360762 | A | G | 5.2619 |

| | | | | | | |
|---|---|---|---|---|---|---|
| *44* | rs440277 | 19 | 45361224 | A | G | -2.8481 |
| *45* | rs411920 | 19 | 45361632 | C | T | -5.0481 |
| *46* | rs365653 | 19 | 45361646 | G | A | -6.2421 |
| *47* | rs418227 | 19 | 45361825 | C | T | 9.2319 |
| *48* | rs417193 | 19 | 45361960 | C | T | 7.3619 |
| *49* | rs2436474 | 19 | 45362269 | T | G | 7.1879 |
| *50* | rs377702 | 19 | 45362667 | A | G | 4.5469 |
| *51* | rs454050 | 19 | 45362809 | A | C | 5.3919 |
| *52* | rs387369 | 19 | 45363254 | A | G | 4.3359 |
| *53* | rs403155 | 19 | 45363299 | T | C | 0.5219 |
| *54* | rs12610257 | 19 | 45363392 | T | C | 6.9379 |
| *55* | rs12978931 | 19 | 45363700 | G | A | 0.3419 |
| *56* | rs384973 | 19 | 45363791 | G | T | -3.9481 |
| *57* | rs411856 | 19 | 45364623 | T | C | 13.2919 |
| *58* | rs395683 | 19 | 45364782 | G | A | 11.3419 |
| *59* | rs395710 | 19 | 45364815 | G | A | -3.5081 |
| *60* | rs555608 | 19 | 45365072 | G | A | 11.5819 |
| *61* | rs2436475 | 19 | 45365248 | C | T | 0.3519 |
| *62* | rs11669109 | 19 | 45365476 | C | T | 4.3919 |
| *63* | rs12980613 | 19 | 45365604 | G | A | 5.0319 |
| *64* | rs12980631 | 19 | 45365641 | C | A | 3.5119 |
| *65* | rs11665829 | 19 | 45365817 | A | G | 4.9019 |
| *66* | rs548011 | 19 | 45365961 | G | A | 5.2969 |
| *67* | rs415637 | 19 | 45366178 | A | G | -2.6181 |
| *68* | rs426555 | 19 | 45366275 | T | C | 0.6019 |
| *69* | rs416116 | 19 | 45366345 | A | G | 1.9219 |
| *70* | rs403729 | 19 | 45366410 | C | G | 4.6019 |
| *71* | rs521629 | 19 | 45366603 | A | G | 2.8419 |
| *72* | rs520566 | 19 | 45366656 | A | G | -6.9781 |
| *73* | rs519825 | 19 | 45366779 | C | T | -1.7181 |
| *74* | rs376938 | 19 | 45366830 | G | A | -2.8831 |
| *75* | rs394353 | 19 | 45367473 | A | G | 7.4019 |
| *76* | rs73050216 | 19 | 45367502 | C | G | 3.6219 |
| *77* | rs8105340 | 19 | 45367777 | C | T | 1.9719 |
| *78* | rs3112439 | 19 | 45367972 | G | C | 9.8359 |
| *79* | rs520283 | 19 | 45368082 | A | C | -3.8181 |
| *80* | rs419010 | 19 | 45368320 | C | T | 4.2579 |
| *81* | rs394221 | 19 | 45368424 | C | T | 11.6919 |
| *82* | rs41290098 | 19 | 45370278 | T | A | 4.0619 |
| *83* | rs384653 | 19 | 45370335 | A | G | 1.6879 |
| *84* | rs511825 | 19 | 45370554 | T | C | 7.7619 |
| *85* | rs565566 | 19 | 45370570 | A | C | 6.6819 |
| *86* | rs511147 | 19 | 45370571 | G | A | -0.4281 |
| *87* | rs564724 | 19 | 45370649 | A | G | 5.6819 |
| *88* | rs510297 | 19 | 45370673 | C | A | 0.2269 |
| *89* | rs12610605 | 19 | 45370838 | A | G | 5.1559 |
| *90* | rs416041 | 19 | 45370854 | G | A | 0.2969 |
| *91* | rs4803766 | 19 | 45371168 | A | G | -0.0281 |

| | | | | | | |
|---|---|---|---|---|---|---|
| *92* | rs390952 | 19 | 45371594 | A | G | -1.9681 |
| *93* | rs58521715 | 19 | 45372129 | T | A | -5.1331 |
| *94* | rs8104483 | 19 | 45372354 | G | T | 1.0719 |
| *95* | rs8104292 | 19 | 45372707 | A | G | 3.6559 |
| *96* | rs404935 | 19 | 45372794 | A | G | -0.8831 |
| *97* | rs559163390 | 19 | 45372849 | C | A | -2.9681 |
| *98* | rs387465 | 19 | 45372867 | G | A | 2.3919 |
| *99* | rs4803767 | 19 | 45372959 | T | C | -0.8781 |
| *100* | rs11879589 | 19 | 45373276 | A | G | 4.3319 |
| *101* | rs3852857 | 19 | 45373539 | C | G | 0.2319 |
| *102* | rs395908 | 19 | 45373565 | A | G | 4.3719 |
| *103* | rs4081918 | 19 | 45373739 | G | A | -1.4531 |
| *104* | rs79074020 | 19 | 45374350 | C | T | 0.4619 |
| *105* | rs519113 | 19 | 45376284 | G | C | 1.4219 |
| *106* | rs393584 | 19 | 45377334 | A | G | 0.5859 |
| *107* | rs2075642 | 19 | 45377467 | A | G | 5.2719 |
| *108* | rs34278513 | 19 | 45378144 | T | C | 3.9119 |
| *109* | rs11665676 | 19 | 45378719 | T | C | -1.6331 |
| *110* | rs387976 | 19 | 45379060 | C | A | 3.4019 |
| *111* | rs3852859 | 19 | 45379309 | C | T | 5.0119 |
| *112* | rs369599 | 19 | 45379336 | T | C | 6.3219 |
| *113* | rs412776 | 19 | 45379516 | A | G | 4.3519 |
| *114* | rs370705 | 19 | 45379638 | T | C | 2.1119 |
| *115* | rs385982 | 19 | 45379682 | C | A | 6.2269 |
| *116* | rs73050293 | 19 | 45379746 | G | A | 0.0419 |
| *117* | rs11667640 | 19 | 45379791 | T | C | 6.7319 |
| *118* | rs419925 | 19 | 45380126 | C | G | 4.3219 |
| *119* | rs421812 | 19 | 45380545 | T | G | 2.4919 |
| *120* | rs3865427 | 19 | 45380961 | A | C | 3.7319 |
| *121* | rs11668861 | 19 | 45380970 | T | G | -1.0941 |
| *122* | rs150639620 | 19 | 45381292 | T | G | 4.1219 |
| *123* | rs3729640 | 19 | 45381917 | T | C | 4.0119 |
| *124* | rs6859 | 19 | 45382034 | A | G | 3.5219 |
| *125* | rs406456 | 19 | 45382717 | G | A | 1.7579 |
| *126* | rs3852860 | 19 | 45382966 | T | C | 3.1719 |
| *127* | rs11669338 | 19 | 45382984 | G | T | -1.5681 |
| *128* | rs11673139 | 19 | 45383037 | T | A | -3.0181 |
| *129* | rs3852861 | 19 | 45383061 | T | G | 5.1619 |
| *130* | rs71352237 | 19 | 45383079 | C | T | 0.0019 |
| *131* | rs34224078 | 19 | 45383115 | G | A | 0.4019 |
| *132* | rs35879138 | 19 | 45383139 | A | T | 1.6119 |
| *133* | rs148303016 | 19 | 45383830 | T | C | -0.8881 |
| *134* | rs11083749 | 19 | 45384105 | T | C | 0.0819 |
| *135* | rs406315 | 19 | 45384116 | G | A | 3.2919 |
| *136* | rs73052307 | 19 | 45384405 | C | T | -1.5941 |
| *137* | rs3745150 | 19 | 45385759 | C | G | -2.5621 |
| *138* | rs149450221 | 19 | 45386040 | A | T | 4.6559 |
| *139* | rs283808 | 19 | 45387034 | C | A | 0.3119 |

| | | | | | | |
|---|---|---|---|---|---|---|
| *140* | rs283809 | 19 | 45387057 | G | A | 0.8669 |
| *141* | rs12972156 | 19 | 45387459 | G | C | -1.0381 |
| *142* | rs12972970 | 19 | 45387596 | A | G | 1.5819 |
| *143* | rs34342646 | 19 | 45388130 | A | G | 3.8219 |
| *144* | rs283810 | 19 | 45388241 | G | T | -3.2381 |
| *145* | rs283811 | 19 | 45388500 | G | A | -0.6331 |
| *146* | rs283813 | 19 | 45389174 | A | T | 0.5719 |
| *147* | rs283814 | 19 | 45389224 | A | G | 1.7119 |
| *148* | rs283815 | 19 | 45390333 | G | A | 3.3619 |
| *149* | rs6857 | 19 | 45392254 | T | C | 2.0819 |
| *150* | rs76692773 | 19 | 45394211 | T | C | 0.4119 |
| *151* | rs71352238 | 19 | 45394336 | C | T | 4.9519 |
| *152* | rs184017 | 19 | 45394969 | G | T | 3.9119 |
| *153* | rs157580 | 19 | 45395266 | G | A | 1.3359 |
| *154* | rs2075649 | 19 | 45395330 | G | A | 5.3669 |
| *155* | rs2075650 | 19 | 45395619 | G | A | 6.4519 |
| *156* | rs157581 | 19 | 45395714 | C | T | 8.5079 |
| *157* | rs34095326 | 19 | 45395844 | A | G | 2.8919 |
| *158* | rs34404554 | 19 | 45395909 | G | C | 0.5019 |
| *159* | rs11556505 | 19 | 45396144 | T | C | 5.1019 |
| *160* | rs157582 | 19 | 45396219 | T | C | 6.6559 |
| *161* | rs59007384 | 19 | 45396665 | T | G | 3.0119 |
| *162* | rs157584 | 19 | 45396899 | C | T | 4.0469 |
| *163* | rs157585 | 19 | 45397512 | C | A | 6.6219 |
| *164* | rs157588 | 19 | 45398264 | T | C | -0.3681 |
| *165* | rs11668327 | 19 | 45398633 | C | G | -1.1281 |
| *166* | rs157590 | 19 | 45398716 | C | A | 2.9719 |
| *167* | rs2238681 | 19 | 45398817 | T | C | 2.7519 |
| *168* | rs8106922 | 19 | 45401666 | G | A | 5.6559 |
| *169* | rs55821237 | 19 | 45401782 | C | T | 5.6169 |
| *170* | rs56290633 | 19 | 45401783 | T | C | 2.9379 |
| *171* | rs34878901 | 19 | 45402477 | T | C | 2.8519 |
| *172* | rs35568738 | 19 | 45402718 | C | G | -0.2031 |
| *173* | rs1160985 | 19 | 45403412 | T | C | 3.4769 |
| *174* | rs760136 | 19 | 45403858 | G | A | 4.5859 |
| *175* | rs1160984 | 19 | 45403924 | T | C | 4.0319 |
| *176* | rs741780 | 19 | 45404431 | C | T | 1.8669 |
| *177* | rs405697 | 19 | 45404691 | A | G | -5.1881 |
| *178* | rs1038025 | 19 | 45404972 | C | T | 2.5819 |
| *179* | rs1038026 | 19 | 45405062 | G | A | 2.6219 |
| *180* | rs1305062 | 19 | 45405521 | C | G | -3.7981 |
| *181* | rs10119 | 19 | 45406673 | A | G | 10.1719 |
| *182* | rs7259620 | 19 | 45407788 | A | G | 1.9419 |
| *183* | rs769446 | 19 | 45408628 | C | T | 12.6519 |
| *184* | rs405509 | 19 | 45408836 | G | T | 5.5619 |
| *185* | rs440446 | 19 | 45409167 | C | G | 4.5419 |
| *186* | rs769449 | 19 | 45410002 | A | G | 1.6319 |
| *187* | rs769450 | 19 | 45410444 | A | G | 0.8619 |

| | | | | | | |
|---|---|---|---|---|---|---|
| *188* | rs429358 | 19 | 45411941 | C | T | 15.2919 |
| *189* | rs7412 | 19 | 45412079 | T | C | -24.2031 |
| *190* | rs1065853 | 19 | 45413233 | G | T | 3.7019 |
| *191* | rs1081106 | 19 | 45413366 | C | T | -4.4281 |
| *192* | rs75627662 | 19 | 45413576 | T | C | 7.8669 |
| *193* | rs439401 | 19 | 45414451 | T | C | 2.9769 |
| *194* | rs10414043 | 19 | 45415713 | A | G | 0.0219 |
| *195* | rs7256200 | 19 | 45415935 | T | G | -0.9281 |
| *196* | rs483082 | 19 | 45416178 | T | G | 3.2419 |
| *197* | rs59325138 | 19 | 45416291 | T | C | 4.8819 |
| *198* | rs584007 | 19 | 45416478 | A | G | -2.2381 |
| *199* | rs438811 | 19 | 45416741 | T | C | 6.2919 |
| *200* | rs5117 | 19 | 45418790 | C | T | 8.6719 |
| *201* | rs3826688 | 19 | 45418961 | T | C | 1.1219 |
| *202* | rs3925681 | 19 | 45421100 | A | G | 1.9119 |
| *203* | rs12721046 | 19 | 45421254 | A | G | 6.2119 |
| *204* | rs12721056 | 19 | 45421744 | T | G | 4.5469 |
| *205* | rs484195 | 19 | 45421877 | A | G | 4.8319 |
| *206* | rs12721051 | 19 | 45422160 | G | C | -3.6881 |
| *207* | rs56131196 | 19 | 45422846 | A | G | -0.2581 |
| *208* | rs4420638 | 19 | 45422946 | G | A | 1.2519 |
| *209* | rs78959900 | 19 | 45423636 | A | G | 2.9119 |
| *210* | rs814573 | 19 | 45424351 | A | T | 5.0859 |
| *211* | rs157592 | 19 | 45424514 | A | C | 7.5219 |

A1, effect allele; A2, reference allele; BP, base position; Chr, chromosome; rsID, dbSNP identifier.

*Table S3. Mean and SE of Evo2 scores and APOE expression levels across the five major ancestry groups from array expression data.*

| | Evo2 score | | APOE expression | |
|---|---|---|---|---|
| **Ancestry** | **mean** | **SE** | **mean** | **SE** |
| **AFR** | -3.80E-04 | 3.01E-05 | -0.321 | 0.067 |
| **AMR** | -4.16E-04 | 5.88E-05 | -0.296 | 0.124 |
| **EAS** | -2.23E-04 | 1.81E-05 | 0.226 | 0.081 |
| **EUR** | -2.70E-04 | 3.28E-05 | 0.213 | 0.097 |
| **SAS** | -3.08E-04 | 2.89E-05 | 0.153 | 0.105 |

AFR, African; AMR, American; EAS, East Asian; EUR, European; SAS, South Asian; SE, standard error.

*Table S4. Association of Evo2 haplotype scores with APOE expression in Pangenome data.*

| | **Beta** | **SE** | **df** | **t value** | **Pr(>\|t\|)** |
|---|---|---|---|---|---|
| **leading haplotype** | 393.08 | 885.22 | 31.00 | 0.44 | 0.660 |
| **alternative haplotype** | 1820.77 | 726.54 | 30.38 | 2.51 | **0.018** |

Linear mix model; n=33. Beta, effect size; df, degree of freedom; SE, standard error.

*Table S5. Correlation of Evo2 haplotype scores with local ancestry burden estimated using RFMix2 in Pangenome data.*

| CRF anchors coordinates (GRCh38) | Value | African | American | East Asian | European | South Asian |
|---|---|---|---|---|---|---|
| 19:44847498 | Spearman r | -0.125 | 0.036 | 0.048 | -0.102 | -0.098 |
| | p-value | **0.0078** | 0.4490 | 0.3070 | **0.0289** | **0.0369** |
| 19:44856449 | Spearman r | -0.115 | -0.015 | 0.119 | -0.063 | -0.117 |
| | p-value | **0.0144** | 0.7490 | **0.0114** | 0.1820 | **0.0125** |
| 19:44867581 | Spearman r | -0.120 | -0.036 | 0.130 | -0.042 | -0.098 |
| | p-value | **0.0102** | 0.4430 | **0.0055** | 0.3730 | **0.0360** |
| 19:44869097 | Spearman r | -0.119 | -0.069 | 0.140 | -0.026 | -0.092 |
| | p-value | **0.0114** | 0.1410 | **0.0028** | 0.5860 | **0.0488** |
| 19:44876534 | Spearman r | -0.137 | -0.120 | 0.164 | -0.068 | -0.074 |
| | p-value | **0.0035** | **0.0104** | **0.0004** | 0.1490 | 0.1140 |
| 19:44880326 | Spearman r | -0.136 | -0.123 | 0.157 | -0.020 | -0.071 |
| | p-value | **0.0035** | **0.0086** | **0.0008** | 0.6740 | 0.1320 |
| 19:44892962 | Spearman r | -0.113 | -0.082 | 0.144 | -0.014 | -0.109 |
| | p-value | **0.0158** | 0.0804 | **0.0021** | 0.7700 | **0.0201** |
| 19:44904531 | Spearman r | -0.124 | -0.055 | 0.149 | -0.014 | -0.107 |
| | p-value | **0.0082** | 0.2440 | **0.0015** | 0.7740 | **0.0222** |
| 19:44911135 | Spearman r | -0.127 | -0.056 | 0.151 | -0.012 | -0.109 |
| | p-value | **0.0066** | 0.2380 | **0.0012** | 0.8000 | **0.0204** |
| 19:44919589 | Spearman r | -0.133 | -0.057 | 0.165 | -0.004 | -0.116 |
| | p-value | **0.0045** | 0.2270 | **0.0004** | 0.9370 | **0.0130** |

Bold indicates p-value < 0.05. CRF, conditional-random-field.

*Table S6. Mean and SE of Evo2 haplotype scores for APOE-ε4 across the five major ancestry groups in Pangenome data.*

| | **Evo2 score (*APOE*-ε4 tagging haplotype)** | | ***APOE*-ε4 odds ratios** | |
|---|---|---|---|---|
| **Ancestry** | **mean** | **SE** | **mean** | **SE** |
| **AFR** | -2.52E-04 | 6.05E-05 | 2.18 | 0.15 |
| **AMR** | -2.29E-04 | 1.12E-04 | 1.90 | 0.14 |
| **EAS** | 1.33E-04 | 2.99E-05 | 4.54 | 0.30 |
| **EUR** | 4.90E-05 | 6.53E-05 | 3.46 | 0.10 |
| **SAS** | -1.24E-04 | 7.00E-05 | | |

AFR, African; AMR, American; EAS, East Asian; EUR, European; SE, standard error. The APOE-ε4 odds ratios for AD was obtained from PMID: 37930705 (*46*).

*Table S7. Association between Evo2 haplotype score and Alzheimer's disease in the ADNI data.*

| AD vs CN | | | | |
|---|---|---|---|---|
| | **Beta** | **SE** | **z_value** | **Pr(>\|z\|)** |
| **ALL** | | | | |
| Leading haplotype | 0.016 | 0.005 | 3.376 | **7.36E-04** |
| Alternative haplotype | 0.004 | 0.004 | 1.169 | 2.42E-01 |
| ***APOE*-ε4 (0 copies)** | | | | |
| Leading haplotype | 0.014 | 0.007 | 2.044 | **4.09E-02** |
| Alternative haplotype | -0.005 | 0.005 | -1.029 | 3.04E-01 |
| ***APOE*-ε4 (1 copy)** | | | | |
| Leading haplotype | -0.029 | 0.019 | -1.538 | 1.24E-01 |
| Alternative haplotype | 0.003 | 0.006 | 0.447 | 6.55E-01 |
| ***APOE*-ε4 (2 copies)** | | | | |
| Leading haplotype | -0.019 | 0.033 | -0.572 | 5.68E-01 |
| Alternative haplotype | -0.022 | 0.021 | -1.03 | 3.03E-01 |

| AD vs MCI | | | | |
|---|---|---|---|---|
| | **Beta** | **SE** | **z_value** | **Pr(>\|z\|)** |
| **ALL** | | | | |
| Leading haplotype | 0.019 | 0.005 | 4.108 | **4.00E-05** |
| Alternative haplotype | 0.006 | 0.003 | 1.799 | 7.20E-02 |
| ***APOE*-ε4 (0 copies)** | | | | |
| Leading haplotype | 0.024 | 0.007 | 3.544 | **3.94E-04** |
| Alternative haplotype | -0.006 | 0.005 | -1.223 | 2.21E-01 |
| ***APOE*-ε4 (1 copy)** | | | | |
| Leading haplotype | -0.016 | 0.017 | -0.934 | 3.50E-01 |
| Alternative haplotype | 0.013 | 0.006 | 2.374 | **1.76E-02** |
| ***APOE*-ε4 (2 copies)** | | | | |
| Leading haplotype | -0.003 | 0.021 | -0.157 | 8.75E-01 |
| Alternative haplotype | 0.012 | 0.014 | 0.861 | 3.89E-01 |

AD, Alzheimer's disease; Beta, effect size; CN, cognitively normal; MCI, mild cognitive impairment; SE, standard error.

*Table S8. Association of Evo2 haplotype scores with Alzheimer's disease–related endophenotypes in the ADNI data.*

| ALL | | | | |
|---|---|---|---|---|
| | **Beta** | **SE** | **z_value** | **Pr(>\|z\|)** |
| **MMSE** | | | | |
| Leading haplotype | -0.012 | 0.004 | -2.811 | **0.0050** |
| Alternative haplotype | -0.004 | 0.003 | -1.406 | 0.1599 |
| **CDRSB** | | | | |
| Leading haplotype | 0.005 | 0.002 | 2.472 | **0.0135** |
| Alternative haplotype | 0.002 | 0.002 | 1.277 | 0.2017 |
| **ADAS11** | | | | |
| Leading haplotype | 0.025 | 0.009 | 2.637 | **0.0084** |
| Alternative haplotype | 0.014 | 0.007 | 1.903 | 0.0572 |
| **ADAS13** | | | | |
| Leading haplotype | 0.043 | 0.015 | 2.926 | **0.0035** |
| Alternative haplotype | 0.022 | 0.012 | 1.898 | 0.0579 |
| **Whole Brain** | | | | |
| Leading haplotype | -270.900 | 143.900 | -1.883 | 0.0599 |
| Alternative haplotype | 87.980 | 108.800 | 0.809 | 0.4189 |
| **Entorhinal** | | | | |
| Leading haplotype | -3.121 | 1.422 | -2.195 | **0.0283** |
| Alternative haplotype | -1.813 | 1.139 | -1.592 | 0.1117 |
| **Hippocampus** | | | | |
| Leading haplotype | -4.771 | 2.275 | -2.097 | **0.0361** |
| Alternative haplotype | -3.791 | 1.766 | -2.147 | **0.0319** |
| **non-CN** | | | | |
| | **Beta** | **SE** | **z_value** | **Pr(>\|z\|)** |
| **MMSE** | | | | |
| Leading haplotype | -0.009 | 0.006 | -1.714 | 0.0867 |
| Alternative haplotype | -0.005 | 0.004 | -1.286 | 0.1985 |
| **CDRSB** | | | | |
| Leading haplotype | 0.006 | 0.003 | 1.936 | 0.0531 |
| Alternative haplotype | 0.002 | 0.002 | 0.790 | 0.4299 |
| **ADAS11** | | | | |
| Leading haplotype | 0.030 | 0.012 | 2.483 | **0.0131** |
| Alternative haplotype | 0.021 | 0.009 | 2.314 | **0.0208** |
| **ADAS13** | | | | |
| Leading haplotype | 0.047 | 0.018 | 2.606 | **0.0093** |
| Alternative haplotype | 0.031 | 0.013 | 2.341 | **0.0194** |
| **Whole Brain** | | | | |
| Leading haplotype | -362.200 | 171.300 | -2.114 | **0.0347** |
| Alternative haplotype | 74.290 | 127.200 | 0.584 | 0.5592 |

| | | | | |
|---|---|---|---|---|
| **Entorhinal** | | | | |
| Leading haplotype | -2.128 | 1.874 | -1.136 | 0.2563 |
| Alternative haplotype | -2.818 | 1.306 | -2.157 | **0.0312** |
| **Hippocampus** | | | | |
| Leading haplotype | -3.852 | 2.687 | -1.434 | 0.1519 |
| Alternative haplotype | -4.941 | 2.068 | -2.389 | **0.0170** |

ADAS11, 11-item Alzheimer's Disease Assessment Scale–Cognitive Subscale; ADAS13, 13-item Alzheimer's Disease Assessment Scale–Cognitive Subscale; Beta, effect size; CDR-SB, Clinical Dementia Rating–Sum of Boxes; CN, cognitively normal; MMSE, Mini-Mental State Examination; SE, standard error.

*Table S9. Association of Evo2 haplotype score with PET-measured brain amyloid burden in the ADNI data.*

| ALL | | | | |
|---|---|---|---|---|
| | **Beta** | **SE** | **t value** | **Pr(>\|t\|)** |
| Leading haplotype | 0.0033 | 0.0010 | 3.3200 | **9.85E-04** |
| Alternative haplotype | 0.0016 | 0.0007 | 2.1620 | **3.12E-02** |

| MCI | | | | |
|---|---|---|---|---|
| | **Beta** | **SE** | **t value** | **Pr(>\|t\|)** |
| Leading haplotype | 0.0049 | 0.0016 | 2.9720 | **3.46E-03** |
| Alternative haplotype | 0.0016 | 0.0012 | 1.3320 | 1.85E-01 |

Beta, effect size; MCI, mild cognitive impairment; PET, Positron Emission Tomography scan, SE, standard error.

*Table S10. Association of Evo2 haplotype score with Alzheimer's disease in the UK Biobank data.*

| **AD vs. controls (2,572 AD; 117,330 controls)** | | | | |
|---|---|---|---|---|
| | **Beta** | **SE** | **z_value** | **Pr(>\|z\|)** |
| Leading haplotype | 2.84E-02 | 1.91E-03 | 14.818 | **< 2e-16** |
| Alternative haplotype | 7.87E-03 | 1.33E-03 | 5.919 | **3.24E-09** |

AD, Alzheimer's disease; Beta, effect size; SE, standard error.

*Table S11. Association of Evo2 haplotype score with regional grey matter volumes (FAST) in the UK Biobank data (p-value < 0.05).*

| Left Brain (leading haplotype) | | | | | | |
|---|---|---|---|---|---|---|
| **Region** | **Mapped Region** | **Beta** | **SE** | **t_value** | **p_value** | **FDR** |
| I IV Cerebellum | CB | -0.0016 | 0.0005 | -3.5136 | **4.43E-04** | 0.03 |
| V Cerebellum | CB | -0.0013 | 0.0004 | -2.9339 | **3.35E-03** | 0.12 |
| Crus I Cerebellum | CB | -0.0011 | 0.0004 | -2.7317 | **6.31E-03** | 0.14 |
| Frontal Medial Cortex | MFC | 0.0009 | 0.0004 | 2.0578 | **3.96E-02** | 0.31 |
| Ventral Striatum | STR | -0.0011 | 0.0004 | -2.5481 | **1.08E-02** | 0.19 |
| Supracalcarine Cortex | V1C | 0.0014 | 0.0004 | 3.1682 | **1.54E-03** | 0.07 |
| Intracalcarine Cortex | V1C | 0.0012 | 0.0004 | 2.6886 | **7.18E-03** | 0.14 |
| Cuneal Cortex | V1C | 0.0009 | 0.0004 | 1.9953 | **4.60E-02** | 0.31 |
| **Right Brain (leading haplotype)** | | | | | | |
| **Region** | **Mapped Region** | **Beta** | **SE** | **t_value** | **p_value** | **FDR** |
| I IV Cerebellum | CB | -0.0011 | 0.0004 | -2.4327 | **1.50E-02** | 0.19 |
| Crus I Cerebellum | CB | -0.0009 | 0.0004 | -2.1623 | **3.06E-02** | 0.30 |
| V Cerebellum | CB | -0.0009 | 0.0004 | -2.0067 | **4.48E-02** | 0.31 |
| VI Cerebellum | CB | -0.0008 | 0.0004 | -1.9826 | **4.74E-02** | 0.31 |
| Lateral Occipital Cortex superior division | OL | 0.0009 | 0.0004 | 2.4448 | **1.45E-02** | 0.19 |
| Occipital Pole | OL | 0.0009 | 0.0004 | 2.2010 | **2.77E-02** | 0.30 |
| Ventral Striatum | STR | -0.0008 | 0.0004 | -2.0109 | **4.43E-02** | 0.31 |
| Thalamus | THA | -0.0009 | 0.0004 | -2.0126 | **4.42E-02** | 0.31 |
| Middle Temporal Gyrus temporooccipital part | TL | 0.0009 | 0.0004 | 2.2348 | **2.54E-02** | 0.29 |
| Intracalcarine Cortex | V1C | 0.0015 | 0.0004 | 3.5148 | **4.41E-04** | 0.03 |
| Cuneal Cortex | V1C | 0.0011 | 0.0004 | 2.7456 | **6.04E-03** | 0.14 |
| Supracalcarine Cortex | V1C | 0.0010 | 0.0004 | 2.4441 | **1.45E-02** | 0.19 |
| **Left Brain (alternative haplotype)** | | | | | | |
| **Region** | **Mapped Region** | **Beta** | **SE** | **t_value** | **p_value** | **FDR** |
| I IV Cerebellum | CB | 0.0010 | 0.0004 | 2.8467 | **4.42E-03** | 0.11 |
| Crus II Cerebellum | CB | 0.0009 | 0.0003 | 2.7151 | **6.63E-03** | 0.11 |
| V Cerebellum | CB | 0.0008 | 0.0003 | 2.4752 | **1.33E-02** | 0.15 |
| Crus I Cerebellum | CB | 0.0007 | 0.0003 | 2.0528 | **4.01E-02** | 0.25 |
| Precentral Gyrus | M1C | -0.0008 | 0.0003 | -2.7820 | **5.41E-03** | 0.11 |
| Frontal Medial Cortex | MFC | -0.0007 | 0.0003 | -2.1129 | **3.46E-02** | 0.24 |
| Occipital Pole | OL | -0.0007 | 0.0003 | -2.3520 | **1.87E-02** | 0.17 |

| | | | | | | |
|---|---|---|---|---|---|---|
| Central Opercular Cortex | PL | 0.0006 | 0.0003 | 2.1001 | **3.57E-02** | 0.24 |
| Superior Temporal Gyrus posterior division | STC | 0.0009 | 0.0003 | 2.6686 | **7.62E-03** | 0.11 |
| **Right Brain (alternative haplotype)** | | | | | | |
| **Region** | **Mapped Region** | **Beta** | **SE** | **t_value** | **p_value** | **FDR** |
| V Cerebellum | CB | 0.0010 | 0.0003 | 2.8835 | **3.94E-03** | 0.11 |
| VI Cerebellum | CB | 0.0009 | 0.0003 | 2.7772 | **5.49E-03** | 0.11 |
| I IV Cerebellum | CB | 0.0009 | 0.0003 | 2.7325 | **6.29E-03** | 0.11 |
| Crus II Cerebellum | CB | 0.0009 | 0.0003 | 2.6750 | **7.48E-03** | 0.11 |
| X Cerebellum | CB | 0.0009 | 0.0003 | 2.5788 | **9.92E-03** | 0.13 |
| VIIb Cerebellum | CB | 0.0008 | 0.0003 | 2.2372 | **2.53E-02** | 0.20 |
| Inferior Temporal Gyrus posterior division | ITC | 0.0007 | 0.0003 | 2.2596 | **2.39E-02** | 0.20 |
| Frontal Medial Cortex | MFC | -0.0007 | 0.0003 | -2.0879 | **3.68E-02** | 0.24 |
| Frontal Orbital Cortex | OFC | -0.0008 | 0.0003 | -2.9468 | **3.21E-03** | 0.11 |

Beta, effect size; FDR, false discovery rate; SE, standard error. Analyses covered 29,783 participants across 139 brain regions for both leading and alternative haplotypes.

*Table S12. Association of Evo2 haplotype score with susceptibility weighted brain MRI in the UK Biobank data (p-value < 0.05).*

| Magnetic susceptibility | | | | | | |
|---|---|---|---|---|---|---|
| **Region** | **Predictor** | **Beta** | **SE** | **t_value** | **p_value** | **FDR** |
| amygdala (right) | Alternative haplotype | 0.0011 | 0.0004 | 2.6939 | **0.0071** | 0.113 |

| T2star | | | | | | |
|---|---|---|---|---|---|---|
| **Region** | **Predictor** | **Beta** | **SE** | **t_value** | **p_value** | **FDR** |
| thalamus (right) | Leading haplotype | 0.0011 | 0.0005 | 2.2220 | **0.0263** | 0.331 |
| thalamus (left) | Leading haplotype | 0.0010 | 0.0005 | 2.0397 | **0.0414** | 0.331 |

Beta, effect size; FDR, false discovery rate; SE, standard error. Analyses covered 27,749 participants across 16 brain regions for Magnetic susceptibility and T2star imaging data for both leading and alternative haplotypes.

*Table S13. Association of Evo2 haplotype score with Olink plasma proteome in the UK Biobank data (FDR < 0.05).*

| Leading haplotype | | | | | |
|---|---|---|---|---|---|
| **Protein** | **Beta** | **SE** | **t_value** | **p_value** | **FDR** |
| apoe | -0.01783 | 0.00045 | -39.41298 | <E-200 | **<E-200** |
| snap25 | 0.01098 | 0.00047 | 23.44524 | 1.88E-120 | **2.73E-117** |
| ment | 0.00803 | 0.00045 | 17.74819 | 4.09E-70 | **3.97E-67** |
| pla2g7 | 0.00611 | 0.00043 | 14.28480 | 3.67E-46 | **2.68E-43** |
| csnk2a1 | 0.00399 | 0.00049 | 8.18882 | 2.74E-16 | **1.60E-13** |
| fgfbp1 | 0.00258 | 0.00045 | 5.71435 | 1.11E-08 | **5.39E-06** |
| bpifb2 | -0.00266 | 0.00047 | -5.64173 | 1.70E-08 | **6.38E-06** |
| nectin2 | -0.00248 | 0.00044 | -5.63635 | 1.75E-08 | **6.38E-06** |
| brk1 | 0.00253 | 0.00046 | 5.54728 | 2.92E-08 | **9.46E-06** |
| man2b2 | -0.00247 | 0.00047 | -5.21673 | 1.83E-07 | **5.34E-05** |
| palm | 0.00241 | 0.00047 | 5.17067 | 2.35E-07 | **6.22E-05** |
| ern1 | -0.00247 | 0.00048 | -5.14472 | 2.70E-07 | **6.55E-05** |
| pdgfc | -0.00229 | 0.00046 | -4.98137 | 6.34E-07 | **1.42E-04** |
| apof | 0.00202 | 0.00044 | 4.57535 | 4.77E-06 | **9.93E-04** |
| pla2g10 | -0.00197 | 0.00044 | -4.47116 | 7.80E-06 | **1.52E-03** |
| hmox1 | -0.00185 | 0.00044 | -4.16255 | 3.15E-05 | **5.75E-03** |
| cd99l2 | -0.00179 | 0.00044 | -4.03319 | 5.51E-05 | **9.45E-03** |
| gm2a | -0.00174 | 0.00045 | -3.82378 | 1.32E-04 | **2.08E-02** |
| tfpi | 0.00168 | 0.00044 | 3.81653 | 1.36E-04 | **2.08E-02** |
| ctsz | -0.00165 | 0.00044 | -3.78185 | 1.56E-04 | **2.27E-02** |
| lrrc37a2 | -0.00177 | 0.00048 | -3.67465 | 2.39E-04 | **3.26E-02** |
| spon2 | -0.00161 | 0.00044 | -3.66112 | 2.51E-04 | **3.26E-02** |
| creg1 | -0.00165 | 0.00045 | -3.64668 | 2.66E-04 | **3.26E-02** |
| agr2 | -0.00166 | 0.00046 | -3.64405 | 2.69E-04 | **3.26E-02** |
| angptl3 | -0.00145 | 0.00041 | -3.58834 | 3.33E-04 | **3.64E-02** |
| psap | -0.00167 | 0.00046 | -3.58688 | 3.35E-04 | **3.64E-02** |
| glb1 | -0.00156 | 0.00044 | -3.58507 | 3.37E-04 | **3.64E-02** |
| rp2 | 0.00163 | 0.00046 | 3.55402 | 3.80E-04 | **3.95E-02** |

| Alternative haplotype | | | | | |
|---|---|---|---|---|---|
| **Protein** | **Beta** | **SE** | **t_value** | **p_value** | **FDR** |
| apoe | -0.00419 | 0.00036 | -11.50781 | 1.39E-30 | **4.06E-27** |
| pla2g7 | 0.00170 | 0.00033 | 5.07173 | 3.96E-07 | **5.77E-04** |
| crtam | 0.00169 | 0.00035 | 4.78480 | 1.72E-06 | **1.67E-03** |
| ment | 0.00171 | 0.00036 | 4.70799 | 2.51E-06 | **1.83E-03** |
| serpind1 | -0.00159 | 0.00038 | -4.20795 | 2.58E-05 | **1.32E-02** |
| cxcl16 | -0.00144 | 0.00035 | -4.17433 | 3.00E-05 | **1.32E-02** |
| snap25 | 0.00157 | 0.00038 | 4.16196 | 3.16E-05 | **1.32E-02** |

| | | | | | |
|---|---|---|---|---|---|
| brk1 | 0.00146 | 0.00036 | 4.08330 | 4.45E-05 | **1.62E-02** |
| ceacam16 | 0.00150 | 0.00038 | 3.97923 | 6.93E-05 | **2.24E-02** |
| ldlr | -0.00134 | 0.00035 | -3.80486 | 1.42E-04 | **4.14E-02** |

Beta, effect size; FDR, false discovery rate; SE, standard error.

*Table S14. Phenome-wide association study results for the Evo2 haplotype score in the UK Biobank data (FDR < 0.05).*

| Leading haplotype | | | | | | |
|---|---|---|---|---|---|---|
| **Phecode** | **Disease** | **Beta** | **SE** | **z_value** | **p_value** | **FDR** |
| **290.11** | Alzheimer's disease | 0.0283 | 0.0018 | 15.4080 | 1.45E-53 | 2.24E-50 |
| **290.13** | Senile dementia | 0.0164 | 0.0017 | 9.7596 | 1.68E-22 | 1.30E-19 |
| **272.11** | Hypercholesterolemia | 0.0038 | 0.0004 | 9.1437 | 6.03E-20 | 3.11E-17 |
| **292.4** | Altered mental status | 0.0061 | 0.0011 | 5.3761 | 7.61E-08 | 2.94E-05 |
| **411.4** | Coronary atherosclerosis | 0.0026 | 0.0005 | 4.6589 | 3.18E-06 | 9.84E-04 |
| **290.1** | Dementias | 0.0211 | 0.0049 | 4.3366 | 1.45E-05 | 3.73E-03 |
| **272.1** | Hyperlipidemia | 0.0036 | 0.0009 | 4.1053 | 4.04E-05 | 8.93E-03 |
| **411.3** | Angina pectoris | 0.0025 | 0.0006 | 4.0448 | 5.24E-05 | 1.01E-02 |
| **694.2** | Other dyschromia | 0.0091 | 0.0025 | 3.6919 | 2.23E-04 | 3.83E-02 |

| Alternative haplotype | | | | | | |
|---|---|---|---|---|---|---|
| **Phecode** | | **Beta** | **SE** | **z_value** | **p_value** | **FDR** |
| **290.13** | Senile dementia | 0.0079 | 0.0012 | 6.5332 | 6.44E-11 | 9.96E-08 |
| **290.11** | Alzheimer's disease | 0.0078 | 0.0013 | 6.1179 | 9.48E-10 | 7.33E-07 |

Beta, effect size; FDR, false discovery rate; SE, standard error. ICD-10 codes were mapped to Phecodes using Phecode Map 1.2 from the PheWAS Resources repository (https://phewascatalog.org/).

5. HPRC Data Explorer. https://data.humanpangenome.org/assemblies.

6. W.-W. Liao, M. Asri, J. Ebler, D. Doerr, M. Haukness, G. Hickey, S. Lu, J. K. Lucas, J. Monlong, H. J. Abel, S. Buonaiuto, X. H. Chang, H. Cheng, J. Chu, V. Colonna, J. M. Eizenga, X. Feng, C. Fischer, R. S. Fulton, S. Garg, C. Groza, A. Guarracino, W. T. Harvey, S. Heumos, K. Howe, M. Jain, T.-Y. Lu, C. Markello, F. J. Martin, M. W. Mitchell, K. M. Munson, M. N. Mwaniki, A. M. Novak, H. E. Olsen, T. Pesout, D. Porubsky, P. Prins, J. A. Sibbesen, J. Sirén, C. Tomlinson, F. Villani, M. R. Vollger, L. L. Antonacci-Fulton, G. Baid, C. A. Baker, A. Belyaeva, K. Billis, A. Carroll, P.-C. Chang, S. Cody, D. E. Cook, R. M. Cook-Deegan, O. E. Cornejo, M. Diekhans, P. Ebert, S. Fairley, O. Fedrigo, A. L. Felsenfeld, G. Formenti, A. Frankish, Y. Gao, N. A. Garrison, C. G. Giron, R. E. Green, L. Haggerty, K. Hoekzema, T. Hourlier, H. P. Ji, E. E. Kenny, B. A. Koenig, A. Kolesnikov, J. O. Korbel, J. Kordosky, S. Koren, H. Lee, A. P. Lewis, H. Magalhães, S. Marco-Sola, P. Marijon, A. McCartney, J. McDaniel, J. Mountcastle, M. Nattestad, S. Nurk, N. D. Olson, A. B. Popejoy, D. Puiu, M. Rautiainen, A. A. Regier, A. Rhie, S. Sacco, A. D. Sanders, V. A. Schneider, B. I. Schultz, K. Shafin, M. W. Smith, H. J. Sofia, A. N. Abou Tayoun, F. Thibaud-Nissen, F. F. Tricomi, J. Wagner, B. Walenz, J. M. D. Wood, A. V. Zimin, G. Bourque, M. J. P. Chaisson, P. Flicek, A. M. Phillippy, J. M. Zook, E. E. Eichler, D. Haussler, T. Wang, E. D. Jarvis, K. H. Miga, E. Garrison, T. Marschall, I. M. Hall, H. Li, B. Paten, A draft human pangenome reference. *Nature* **617**, 312–324 (2023).

7. IGSR | samples. https://www.internationalgenome.org/data-portal/sample.

8. S. Fairley, E. Lowy-Gallego, E. Perry, P. Flicek, The International Genome Sample Resource (IGSR) collection of open human genomic variation resources. *Nucleic Acids Research* **48**, D941–D947 (2020).

9. RNAseq | 1000 Genomes. https://www.internationalgenome.org/category/rnaseq/.

20. F. Abascal, R. Acosta, N. J. Addleman, J. Adrian, V. Afzal, R. Ai, B. Aken, J. A. Akiyama, O. A. Jammal, H. Amrhein, S. M. Anderson, G. R. Andrews, I. Antoshechkin, K. G. Ardlie, J. Armstrong, M. Astley, B. Banerjee, A. A. Barkal, I. H. A. Barnes, I. Barozzi, D. Barrell, G. Barson, D. Bates, U. K. Baymuradov, C. Bazile, M. A. Beer, S. Beik, M. A. Bender, R. Bennett, L. P. B. Bouvrette, B. E. Bernstein, A. Berry, A. Bhaskar, A. Bignell, S. M. Blue, D. M. Bodine, C. Boix, N. Boley, T. Borrman, B. Borsari, A. P. Boyle, L. A. Brandsmeier, A. Breschi, E. H. Bresnick, J. A. Brooks, M. Buckley, C. B. Burge, R. Byron, E. Cahill, L. Cai, L. Cao, M. Carty, R. G. Castanon, A. Castillo, H. Chaib, E. T. Chan, D. R. Chee, S. Chee, H. Chen, H. Chen, J.-Y. Chen, S. Chen, J. M. Cherry, S. B. Chhetri, J. S. Choudhary, J. Chrast, D. Chung, D. Clarke, N. A. L. Cody, C. J. Coppola, J. Coursen, A. M. D'Ippolito, S. Dalton, C. Danyko, C. Davidson, J. Davila-Velderrain, C. A. Davis, J. Dekker, A. Deran, G. DeSalvo, G. Despacio-Reyes, C. N. Dewey, D. E. Dickel, M. Diegel, M. Diekhans, V. Dileep, B. Ding, S. Djebali, A. Dobin, D. Dominguez, S. Donaldson, J. Drenkow, T. R. Dreszer, Y. Drier, M. O. Duff, D. Dunn, C. Eastman, J. R. Ecker, M. D. Edwards, N. El-Ali, S. I. Elhajjajy, K. Elkins, A. Emili, C. B. Epstein, R. C. Evans, I. Ezkurdia, K. Fan, P. J. Farnham, N. P. Farrell, E. A. Feingold, A.-M. Ferreira, K. Fisher-Aylor, S. Fitzgerald, P. Flicek, C. S. Foo, K. Fortier, A. Frankish, P. Freese, S. Fu, X.-D. Fu, Y. Fu, Y. Fukuda-Yuzawa, M. Fulciniti, A. P. W. Funnell, I. Gabdank, T. Galeev, M. Gao, C. G. Giron, T. H. Garvin, C. A. Gelboin-Burkhart, G. Georgolopoulos, M. B. Gerstein, B. M. Giardine, D. K. Gifford, D. M. Gilbert, D. A. Gilchrist, S. Gillespie, T. R. Gingeras, P. Gong, A. Gonzalez, J. M. Gonzalez, P. Good, A. Goren, D. U. Gorkin, B. R. Graveley, M. Gray, J. F. Greenblatt, E. Griffiths, M. T. Groudine, F. Grubert, M. Gu, R. Guigó, H. Guo, Y. Guo, Y. Guo, G. Gursoy, M. Gutierrez-Arcelus, J. Halow, R. C. Hardison, M. Hardy, M. Hariharan, A. Harmanci, A. Harrington, J. L. Harrow, T. B. Hashimoto, R. D. Hasz, M. Hatan, E. Haugen, J. E. Hayes, P. He, Y. He, N. Heidari, D. Hendrickson, E. F. Heuston, J. A. Hilton, B. C. Hitz, A. Hochman, C. Holgren, L. Hou, S. Hou, Y.-H. E. Hsiao, S. Hsu, H. Huang, T. J. Hubbard, J. Huey, T. R. Hughes, T. Hunt, S. Ibarrientos, R. Issner, M. Iwata, O. Izuogu, T. Jaakkola, N. Jameel, C. Jansen, L. Jiang, P. Jiang, A. Johnson, R. Johnson, I. Jungreis, M. Kadaba, M. Kasowski, M. Kasparian, M. Kato, R. Kaul, T. Kawli, M. Kay, J. C. Keen, S. Keles, C. A. Keller, D. Kelley, M. Kellis, P. Kheradpour, D. S. Kim, A. Kirilusha, R. J. Klein, B. Knoechel, S. Kuan, M. J. Kulik, S. Kumar, A. Kundaje, T. Kutyavin, J. Lagarde, B. R. Lajoie, N. J. Lambert, J. Lazar, A. Y. Lee, D. Lee, E. Lee, J. W. Lee, K. Lee, C. S. Leslie, S. Levy, B. Li, H. Li, N. Li, S. Li, X. Li, Y. I. Li, Y. Li, Y. Li, Y. Li, J. Lian, M. W. Libbrecht, S. Lin, Y. Lin, D. Liu, J. Liu, P. Liu, T. Liu, X. S. Liu, Y. Liu, Y. Liu, M. Long, S. Lou, J. Loveland, A. Lu, Y. Lu, E. Lécuyer, L. Ma, M. Mackiewicz, B. J. Mannion, M. Mannstadt, D. Manthravadi, G. K. Marinov, F. J. Martin, E. Mattei, K. McCue, M. McEown, G. McVicker, S. K. Meadows, A. Meissner, E. M. Mendenhall, C. L. Messer, W. Meuleman, C. Meyer, S. Miller, M. G. Milton, T. Mishra, D. E. Moore, H. M. Moore, J. E. Moore, S. H. Moore, J. Moran, A. Mortazavi, J. M. Mudge, N. Munshi, R. Murad, R. M. Myers, V. Nandakumar, P. Nandi, A. M. Narasimha, A. K. Narayanan, H. Naughton, F. C. P. Navarro, P. Navas, J. Nazarovs, J. Nelson, S. Neph, F. J. Neri, The ENCODE Project Consortium, Expanded encyclopaedias of DNA elements in the human and mouse genomes. *Nature* **583**, 699–710 (2020).

21. H. Li, Minimap2: pairwise alignment for nucleotide sequences. *Bioinformatics* **34**, 3094–3100 (2018).

22. H. Li, A statistical framework for SNP calling, mutation discovery, association mapping and population genetical parameter estimation from sequencing data. *Bioinformatics* **27**, 2987–2993 (2011).

23. B. K. Maples, S. Gravel, E. E. Kenny, C. D. Bustamante, RFMix: A Discriminative Modeling Approach for Rapid and Robust Local-Ancestry Inference. *The American Journal of Human Genetics* **93**, 278–288 (2013).

24. A. Auton, G. R. Abecasis, D. M. Altshuler, R. M. Durbin, G. R. Abecasis, D. R. Bentley, A. Chakravarti, A. G. Clark, P. Donnelly, E. E. Eichler, P. Flicek, S. B. Gabriel, R. A. Gibbs, E. D. Green, M. E. Hurles, B. M. Knoppers, J. O. Korbel, E. S. Lander, C. Lee, H. Lehrach, E. R. Mardis, G. T. Marth, G. A. McVean, D. A. Nickerson, J. P. Schmidt, S. T. Sherry, J. Wang, R. K. Wilson, R. A. Gibbs, E. Boerwinkle, H. Doddapaneni, Y. Han, V. Korchina, C. Kovar, S. Lee, D. Muzny, J. G. Reid, Y. Zhu, J. Wang, Y. Chang, Q. Feng, X. Fang, X. Guo, M. Jian, H. Jiang, X. Jin, T. Lan, G. Li, J. Li, Y. Li, S. Liu, X. Liu, Y. Lu, X. Ma, M. Tang, B. Wang, G. Wang, H. Wu, R. Wu, X. Xu, Y. Yin, D. Zhang, W. Zhang, J. Zhao, M. Zhao, X. Zheng, E. S. Lander, D. M. Altshuler, S. B. Gabriel, N. Gupta, N. Gharani, L. H. Toji, N. P. Gerry, A. M. Resch, P. Flicek, J. Barker, L. Clarke, L. Gil, S. E. Hunt, G. Kelman, E. Kulesha, R. Leinonen, W. M. McLaren, R. Radhakrishnan, A. Roa, D. Smirnov, R. E. Smith, I. Streeter, A. Thormann, I. Toneva, B. Vaughan, X. Zheng-Bradley, D. R. Bentley, R. Grocock, S. Humphray, T. James, Z. Kingsbury, H. Lehrach, R. Sudbrak, M. W. Albrecht, V. S. Amstislavskiy, T. A. Borodina, M. Lienhard, F. Mertes, M. Sultan, B. Timmermann, M.-L. Yaspo, E. R. Mardis, R. K. Wilson, L. Fulton, R. Fulton, S. T. Sherry, V. Ananiev, Z. Belaia, D. Beloslyudtsev, N. Bouk, C. Chen, D. Church, R. Cohen, C. Cook, J. Garner, T. Hefferon, M. Kimelman, C. Liu, J. Lopez, P. Meric, C. O'Sullivan, Y. Ostapchuk, L. Phan, S. Ponomarov, V. Schneider, E. Shekhtman, K. Sirotkin, D. Slotta, H. Zhang, G. A. McVean, R. M. Durbin, S. Balasubramaniam, J. Burton, P. Danecek, T. M. Keane, A. Kolb-Kokocinski, S. McCarthy, J. Stalker, M. Quail, J. P. Schmidt, C. J. Davies, J. Gollub, T. Webster, B. Wong, Y. Zhan, A. Auton, C. L. Campbell, Y. Kong, A. Marcketta, R. A. Gibbs, F. Yu, L. Antunes, M. Bainbridge, D. Muzny, A. Sabo, Z. Huang, J. Wang, L. J. M. Coin, L. Fang, X. Guo, X. Jin, G. Li, Q. Li, Y. Li, Z. Li, H. Lin, B. Liu, R. Luo, H. Shao, Y. Xie, C. Ye, C. Yu, F. Zhang, H. Zheng, H. Zhu, C. Alkan, E. Dal, F. Kahveci, G. T. Marth, E. P. Garrison, D. Kural, W.-P. Lee, W. Fung Leong, M. Stromberg, A. N. Ward, J. Wu, M. Zhang, M. J. Daly, M. A. DePristo, R. E. Handsaker, D. M. Altshuler, E. Banks, G. Bhatia, G. del Angel, S. B. Gabriel, G. Genovese, N. Gupta, H. Li, S. Kashin, E. S. Lander, S. A. McCarroll, J. C. Nemesh, R. E. Poplin, S. C. Yoon, J. Lihm, V. Makarov, A. G. Clark, S. Gottipati, A. Keinan, J. L. Rodriguez-Flores, J. O. Korbel, T. Rausch, M. H. Fritz, A. M. Stütz, P. Flicek, K. Beal, L. Clarke, A. Datta, J. Herrero, W. M. McLaren, G. R. S. Ritchie, R. E. Smith, D. Zerbino, X. Zheng-Bradley, P. C. Sabeti, I. Shlyakhter, S. F. Schaffner, J. Vitti, D. N. Cooper, E. V. Ball, P. D. Stenson, D. R. Bentley, B. Barnes, M. Bauer, R. Keira Cheetham, A. Cox, M. Eberle, S. Humphray, S. Kahn, L. Murray, J. Peden, R. Shaw, E. E. Kenny, M. A. Batzer, M. K. Konkel, J. A. Walker, D. G. MacArthur, M. Lek, R. Sudbrak, V. S. Amstislavskiy, R. Herwig, E. R. Mardis, L. Ding, D. C. Koboldt, D. Larson, K. Ye, S. Gravel, The 1000 Genomes Project Consortium, Corresponding authors, Steering committee, Production group, Baylor College of Medicine, BGI-Shenzhen, Broad Institute of MIT and Harvard, Coriell Institute for Medical Research, E. B. I. European Molecular Biology Laboratory, Illumina, Max Planck Institute for Molecular Genetics, McDonnell Genome Institute at Washington University, US National Institutes of Health, University of Oxford, Wellcome Trust

Sanger Institute, Analysis group, Affymetrix, Albert Einstein College of Medicine, Bilkent University, Boston College, Cold Spring Harbor Laboratory, Cornell University, European Molecular Biology Laboratory, Harvard University, Human Gene Mutation Database, Icahn School of Medicine at Mount Sinai, Louisiana State University, Massachusetts General Hospital, McGill University, N. National Eye Institute, A global reference for human genetic variation. *Nature* **526**, 68–74 (2015).

25. ).

26. P.-C. Bürkner, brms: An R Package for Bayesian Multilevel Models Using Stan. *Journal of Statistical Software* **80**, 1–28 (2017).

27. A. Kuznetsova, P. B. Brockhoff, R. H. B. Christensen, lmerTest Package: Tests in Linear Mixed Effects Models. *Journal of Statistical Software* **82**, 1–26 (2017).

28. A. S. Hinrichs, D. Karolchik, R. Baertsch, G. P. Barber, G. Bejerano, H. Clawson, M. Diekhans, T. S. Furey, R. A. Harte, F. Hsu, J. Hillman-Jackson, R. M. Kuhn, J. S. Pedersen, A. Pohl, B. J. Raney, K. R. Rosenbloom, A. Siepel, K. E. Smith, C. W. Sugnet, A. Sultan-Qurraie, D. J. Thomas, H. Trumbower, R. J. Weber, M. Weirauch, A. S. Zweig, D. Haussler, W. J. Kent, The UCSC Genome Browser Database: update 2006. *Nucleic Acids Research* **34**, D590–D598 (2006).

29. S. Das, L. Forer, S. Schönherr, C. Sidore, A. E. Locke, A. Kwong, S. I. Vrieze, E. Y. Chew, S. Levy, M. McGue, D. Schlessinger, D. Stambolian, P.-R. Loh, W. G. Iacono, A. Swaroop, L. J. Scott, F. Cucca, F. Kronenberg, M. Boehnke, G. R. Abecasis, C. Fuchsberger, Next-generation genotype imputation service and methods. *Nat Genet* **48**, 1284–1287 (2016).

30. S. McCarthy, S. Das, W. Kretzschmar, O. Delaneau, A. R. Wood, A. Teumer, H. M. Kang, C. Fuchsberger, P. Danecek, K. Sharp, Y. Luo, C. Sidore, A. Kwong, N. Timpson, S. Koskinen, S. Vrieze, L. J. Scott, H. Zhang, A. Mahajan, J. Veldink, U. Peters, C. Pato, C. M. van Duijn, C. E. Gillies, I. Gandin, M. Mezzavilla, A. Gilly, M. Cocca, M. Traglia, A. Angius, J. C. Barrett, D. Boomsma, K. Branham, G. Breen, C. M. Brummett, F. Busonero, H. Campbell, A. Chan, S. Chen, E. Chew, F. S. Collins, L. J. Corbin, G. D. Smith, G. Dedoussis, M. Dorr, A.-E. Farmaki, L. Ferrucci, L. Forer, R. M. Fraser, S. Gabriel, S. Levy, L. Groop, T. Harrison, A. Hattersley, O. L. Holmen, K. Hveem, M. Kretzler, J. C. Lee, M. McGue, T. Meitinger, D. Melzer, J. L. Min, K. L. Mohlke, J. B. Vincent, M. Nauck, D. Nickerson, A. Palotie, M. Pato, N. Pirastu, M. McInnis, J. B. Richards, C. Sala, V. Salomaa, D. Schlessinger, S. Schoenherr, P. E. Slagboom, K. Small, T. Spector, D. Stambolian, M. Tuke, J. Tuomilehto, L. H. Van den Berg, W. Van Rheenen, U. Volker, C. Wijmenga, D. Toniolo, E. Zeggini, P. Gasparini, M. G. Sampson, J. F. Wilson, T. Frayling, P. I. W. de Bakker, M. A. Swertz, S. McCarroll, C. Kooperberg, A. Dekker, D. Altshuler, C. Willer, W. Iacono, S. Ripatti, N. Soranzo, K. Walter, A. Swaroop, F. Cucca, C. A. Anderson, R. M. Myers, M. Boehnke, M. I. McCarthy, R. Durbin, G. Abecasis, J. Marchini, the Haplotype Reference Consortium, A reference panel of 64,976 haplotypes for genotype imputation. *Nat Genet* **48**, 1279–1283 (2016).

31. V. Todorov, P. Filzmoser, An Object-Oriented Framework for Robust Multivariate Analysis. *Journal of Statistical Software* **32**, 1–47 (2010).

32. B. L. Browning, S. R. Browning, Genotype Imputation with Millions of Reference Samples. *The American Journal of Human Genetics* **98**, 116–126 (2016).

33. A. Rakowski, R. Monti, C. Lippert, TransferGWAS of T1-weighted brain MRI data from UK Biobank. *PLOS Genetics* **20**, e1011332 (2024).

34. C. Wang, A. B. Martins-Bach, F. Alfaro-Almagro, G. Douaud, J. C. Klein, A. Llera, C. Fiscone, R. Bowtell, L. T. Elliott, S. M. Smith, B. C. Tendler, K. L. Miller, Phenotypic and genetic associations of quantitative magnetic susceptibility in UK Biobank brain imaging. *Nature Neuroscience* **25**, 818–831 (2022).

35. G. H. Eldjarn, E. Ferkingstad, S. H. Lund, H. Helgason, O. Th. Magnusson, K. Gunnarsdottir, T. A. Olafsdottir, B. V. Halldorsson, P. I. Olason, F. Zink, S. A. Gudjonsson, G. Sveinbjornsson, M. I. Magnusson, A. Helgason, A. Oddsson, G. H. Halldorsson, M. K. Magnusson, S. Saevarsdottir, T. Eiriksdottir, G. Masson, H. Stefansson, I. Jonsdottir, H. Holm, T. Rafnar, P. Melsted, J. Saemundsdottir, G. L. Norddahl, G. Thorleifsson, M. O. Ulfarsson, D. F. Gudbjartsson, U. Thorsteinsdottir, P. Sulem, K. Stefansson, Large-scale plasma proteomics comparisons through genetics and disease associations. *Nature* **622**, 348–358 (2023).

36. L. Kolberg, U. Raudvere, I. Kuzmin, J. Vilo, H. Peterson, gprofiler2 -- an R package for gene list functional enrichment analysis and namespace conversion toolset g:Profiler. F1000Research 9:709 [Preprint] (2020). https://doi.org/10.12688/f1000research.24956.2.

37. J. C. Denny, L. Bastarache, M. D. Ritchie, R. J. Carroll, R. Zink, J. D. Mosley, J. R. Field, J. M. Pulley, A. H. Ramirez, E. Bowton, M. A. Basford, D. S. Carrell, P. L. Peissig, A. N. Kho, J. A. Pacheco, L. V. Rasmussen, D. R. Crosslin, P. K. Crane, J. Pathak, S. J. Bielinski, S. A. Pendergrass, H. Xu, L. A. Hindorff, R. Li, T. A. Manolio, C. G. Chute, R. L. Chisholm, E. B. Larson, G. P. Jarvik, M. H. Brilliant, C. A. McCarty, I. J. Kullo, J. L. Haines, D. C. Crawford, D. R. Masys, D. M. Roden, Systematic comparison of phenome-wide association study of electronic medical record data and genome-wide association study data. *Nature Biotechnology* **31**, 1102–1111 (2013).
